# SARS-CoV-2 Community Transmission During Shelter-in-Place in San Francisco

**DOI:** 10.1101/2020.06.15.20132233

**Authors:** Gabriel Chamie, Carina Marquez, Emily Crawford, James Peng, Maya Petersen, Daniel Schwab, Joshua Schwab, Jackie Martinez, Diane Jones, Douglas Black, Monica Gandhi, Andrew D. Kerkhoff, Vivek Jain, Francesco Sergi, Jon Jacobo, Susana Rojas, Valerie Tulier-Laiwa, Tracy Gallardo-Brown, Ayesha Appa, Charles Chiu, Mary Rodgers, John Hackett, CLIAhub Consortium, Amy Kistler, Samantha Hao, Jack Kamm, David Dynerman, Joshua Batson, Bryan Greenhouse, Joe DeRisi, Diane V. Havlir

## Abstract

**Background:** We characterized SARS-CoV-2 infections in a densely-populated, majority Latinx San Francisco community six-weeks into the city’s shelter-in-place order.

**Methods:** We offered SARS-CoV-2 reverse transcription-PCR and antibody (Abbott ARCHITECT IgG) testing, regardless of symptoms, to all residents (>4 years) and workers in a San Francisco census tract (population: 5,174) at outdoor, community-mobilized events over four days. We estimated SARS-CoV-2 point prevalence (PCR-positive) and cumulative incidence (antibody or PCR-positive) in the census tract and evaluated risk factors for recent (PCR-positive/antibody-negative) versus prior infection (antibody-positive/PCR-negative). SARS-CoV-2 genome recovery and phylogenetics were used to measure viral strain diversity, establish viral lineages present, and estimate number of introductions.

**Results:** We tested 3,953 persons: 40% Latinx; 41% White; 9% Asian/Pacific Islander; and 2% Black. Overall, 2.1% (83/3,871) tested PCR-positive: 95% were Latinx and 52% asymptomatic when tested. 1.7% of residents and 6.0% of workers (non-census tract residents) were PCR-positive. Among 2,598 census tract residents, estimated point prevalence of PCR-positives was 2.3% (95%CI: 1.2-3.8%): 3.9% (95%CI: 2.0-6.4%) among Latinx vs. 0.2% (95%CI: 0.0-0.4%) among non-Latinx persons. Estimated cumulative incidence among residents was 6.1% (95%CI: 4.0-8.6%). Prior infections were 67% Latinx, 16% White, and 17% other ethnicities. Among recent infections, 96% were Latinx. Risk factors for recent infection were Latinx ethnicity, inability to shelter-in-place and maintain income, frontline service work, unemployment, and household income <$50,000/year. Five SARS-CoV-2 phylogenetic lineages were detected.

**Conclusion:** SARS-CoV-2 infections from diverse lineages continued circulating among low-income, Latinx persons unable to work from home and maintain income during San Francisco’s shelter-in-place ordinance.

## Introduction

In early 2020, multiple introductions of SARS-CoV-2 into the United States laid the foundation for the ongoing epidemic that has claimed over 100,000 U.S. lives in less than 6 months.^1^ Some of the earliest clinical cases of COVID-19 were recognized in California,^2^ and the state led the nation in issuing a state-wide shelter-in-place mandate on March 19.^3^ San Francisco declared a local emergency on February 25 and issued a series of increasingly restrictive mandates on sizes of gatherings culminating in a shelter-in place order on March 16. Although peak hospitalization and death rates in San Francisco over the ensuing month were nearly 10-fold lower than hard-hit cities such as New York,^4^ the pattern of disproportionately higher hospitalizations among communities of color was similar.^5^ In San Francisco, 45% of reported COVID-19 cases are among Latinx people, who represent 15% of the city’s population.^6^

Hospitalizations and deaths represent a small fraction of the total SARS-CoV-2 infections in a community.^7^ Estimates of the burden of community SARS-CoV-2 infections from direct measurements have been difficult to obtain and compare because symptomatic testing programs capture only a proportion of cases,^8^ the recognized symptoms associated with COVID-19 expanded over time,^9^ the assays used to identify infection have variable performance characteristics, and easily accessible testing programs are not in place for some of the most highly affected communities. Data on community transmission and ethnic disparities in SARS-CoV-2 infection, as opposed to COVID-19 disease^10^, as well as systematic efforts to determine factors driving these disparities remain limited.^11^

To characterize ongoing community transmission during a citywide shelter-in-place mandate, we offered population-based, universal testing for SARS-CoV-2 infection to all residents of a densely-populated census tract within a majority Latinx community in San Francisco.

## Methods

Unidos en Salud is a longitudinal study to characterize SARS-CoV-2 epidemiology and assess impact of public health measures within a US census tract in San Francisco. Six-weeks into the city’s shelter-in-place ordinance, we offered SARS-CoV-2 reverse transcription-PCR (RT-PCR) and antibody testing, regardless of symptoms, to residents (≥4 years) and people who work but may not reside in the census tract.

### Study Setting and Community Mobilization

U.S. census tract 022901 is a population-dense, 16-square-block (0.1 square-mile) area in San Francisco’s Mission District, with 5,174 residents of whom 58% are Latinx, 34% White/Caucasian, 5% Asian/Pacific Islander, and 1% Black/African American. Median per capita income was $40,420/year in 2018, with 34% of households earning <$50,000/year and 20% earning >$200,000/year.^12^ In partnership with the Latino Task Force for COVID-19, an umbrella organization coordinating local Latinx community-based organizations, we distributed flyers, mobilized the community on local and social media, and offered online and door-to-door pre-registration for testing appointments during the week prior to the testing campaign.

### Testing Campaign

From April 25-28, 2020, we offered outdoor testing at public parks and schools to those who provided an address in the tract or worked in the tract. On April 28, we expanded eligibility to residents of neighboring city blocks, responding to high community demand. One week later, we offered testing to home-bound residents who could not reach campaign sites. During pre-registration, we conducted a brief survey. At the time of testing, we obtained verbal consent for participation and conducted COVID-19 symptom screening. Medical staff performed a fingerstick blood collection (500µL) for antibody testing and an oropharyngeal/mid-turbinate nasal swab for quantitative RT-PCR. Participants could opt out of either test. We contacted all PCR-positive persons to disclose results and perform a clinical assessment. We provided household support via a community-led team for PCR-positive participants and evaluated symptoms among all PCR-positive participants over 2 weeks following testing.

### Laboratory assays

Swabs were collected in DNA/RNA Shield (Zymo Research) to inactivate virus and preserve RNA stability. RT-PCR of viral N and E genes and human RNAse P gene was performed on extracted RNA at a CLIA-certified laboratory operated by UCSF and the Chan Zuckerberg Biohub using a Laboratory Developed Test with a limit of detection of log10 4.5 viral genome copies/mL. SARS-CoV-2-positive RNA samples were subjected to Primal-Seq Nextera XT version 2.0,^13^ using the ARTIC Network V3 primers,^14^ followed by paired-end 2 × 150bp sequencing on an Illumina NovaSeq platform. For antibody testing, the ARCHITECT SARS-CoV-2 IgG Emergency Use Authorization (EUA) assay (Abbott Laboratories, Abbott Park, IL, USA)^15^ was performed on participants’ plasma from the fingerstick collections, which is a research use of the test.

### Study Outcomes

Outcomes included the estimated point prevalence of all PCR-positive infections, recent infections (PCR-positive/antibody-negative) and prior infections (antibody-positive/PCR-negative). Cumulative incidence of infection was defined as any PCR or antibody-positive result. Phylogenetics were used to measure strain diversity.

### Statistical analyses

Proportions were compared using chi-squared tests and medians compared using Wilcoxon rank-sum tests. Cumulative incidence of infection was adjusted for RT-PCR and antibody test characteristics; 95% confidence intervals incorporating uncertainty in test characteristics were based on bootstrap. For census tract residents, we further adjusted for differences in age, sex, and race/ethnicity of participants compared to 2018 census estimates (**Supplementary Methods**). We used multivariate logistic regression for the dependent outcome of PCR-positivity among participants tested. We did not adjust confidence intervals for multiple testing.

### Bioinformatics and genomic analyses

A phylogenetic tree was constructed containing all 123 SARS-CoV-2 genomes from San Francisco County on GISAID^16^ on May 22, 2020 together with the high-quality consensus genomes assembled from this study, using the nextstrain toolkit.^17^ Global clade identification and naming follows the Nextstrain proposal,^18^ and significance of population structure was computed by a permutation test was used for Hudson’s FST (**Supplementary Methods**).^19^

### Ethics Statement & Funding

The UCSF Committee on Human Research determined that the study met criteria for public health surveillance. The study was supported by the Chan Zuckerberg Biohub, UCSF, and a Program for Breakthrough Biomedical Research award. ARCHITECT SARS-CoV-2 test kits were provided by Abbott Laboratories.

## Results

### Testing Uptake and Coverage

From April 25-28, 2020, we tested 3,913 people. From May 6-7, we tested an additional 40 home-bound residents, for a total of 3,953 persons tested. Of all persons tested, 53% were male, and 40% identified as Latinx, 41% White, 9% Asian/Pacific Islander, 2% Black, and 7% other/mixed ethnicity (**Table 1**). Estimated census tract testing coverage of adult residents (age >20 years) was 60%.

**Table 1.**
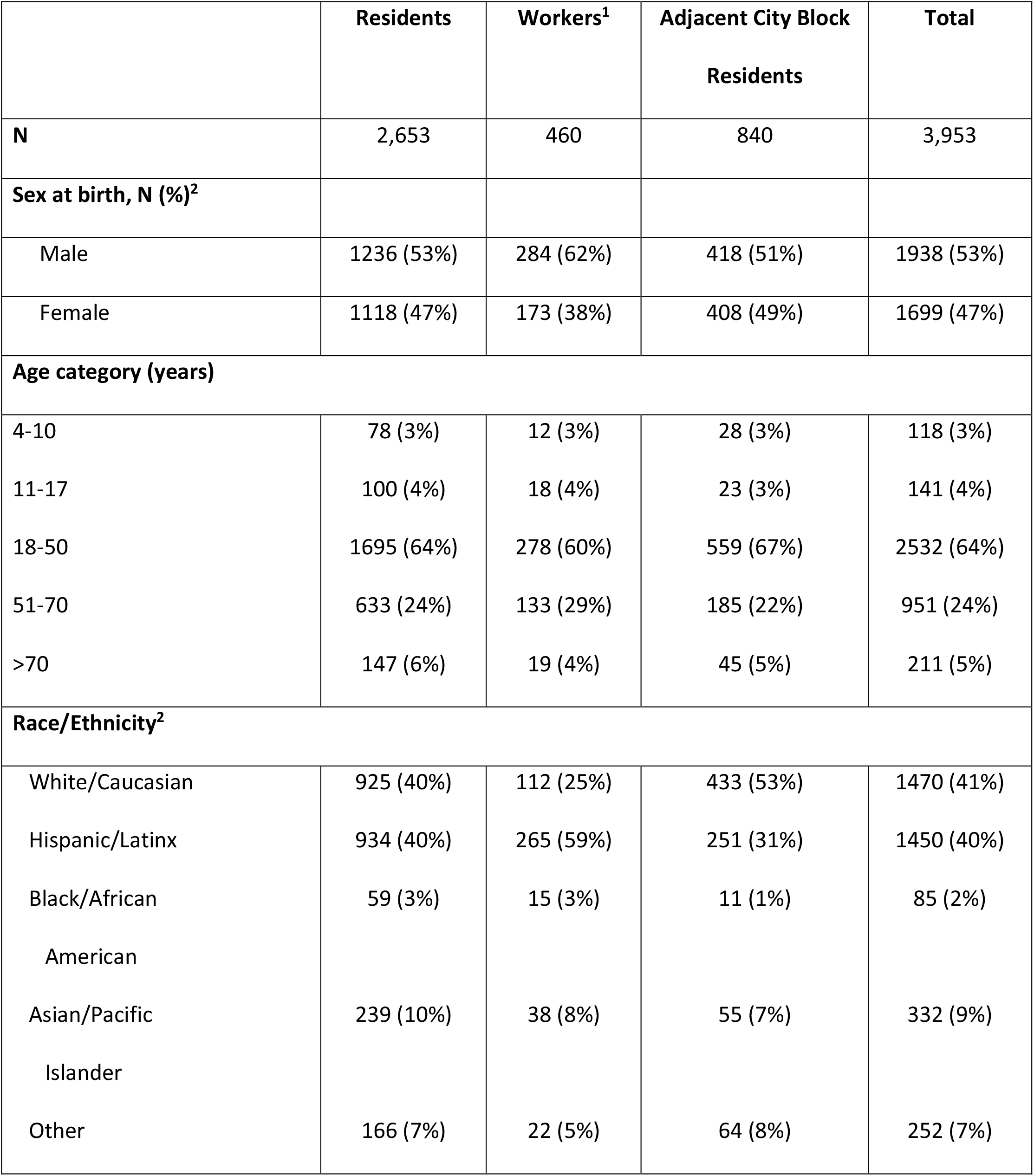
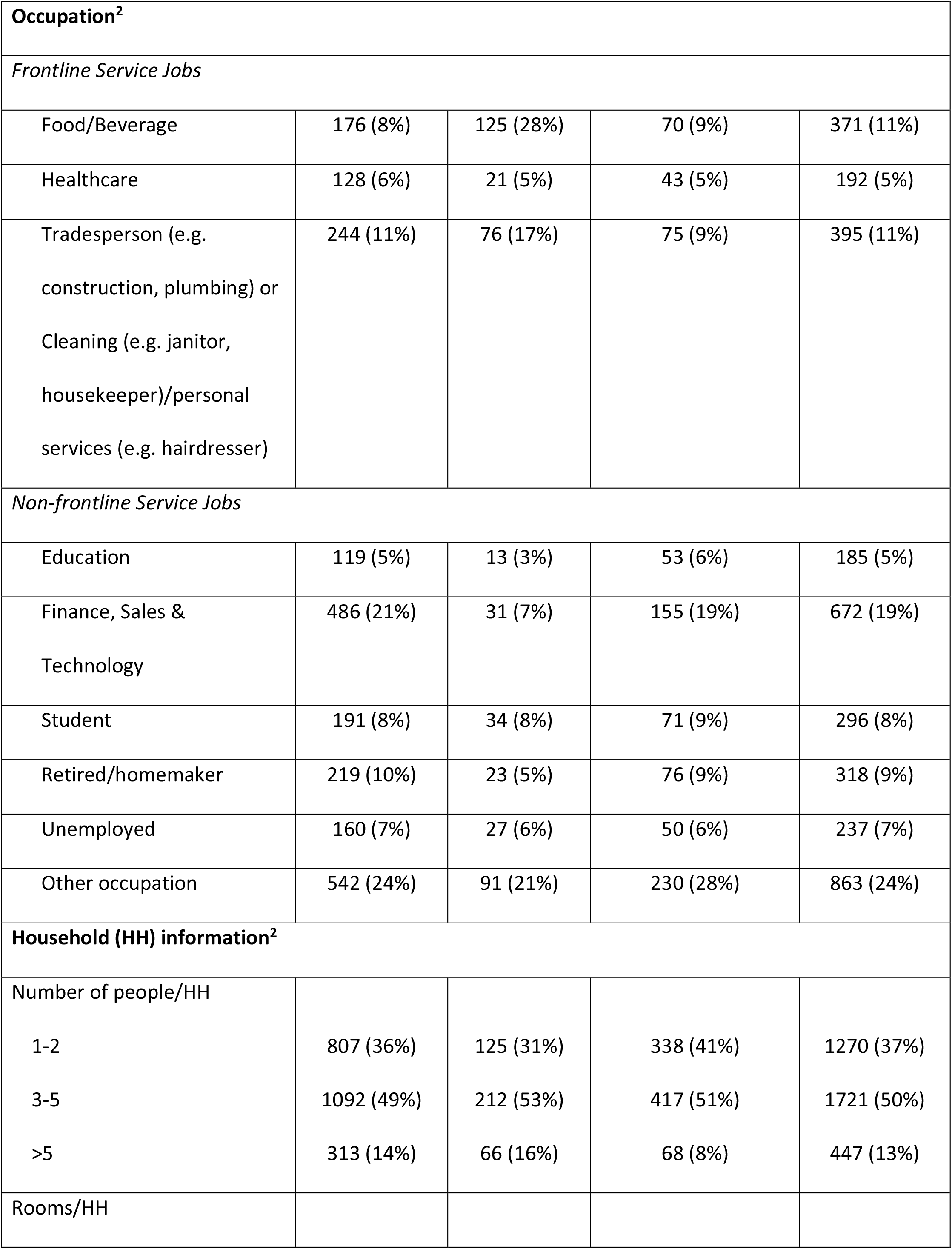
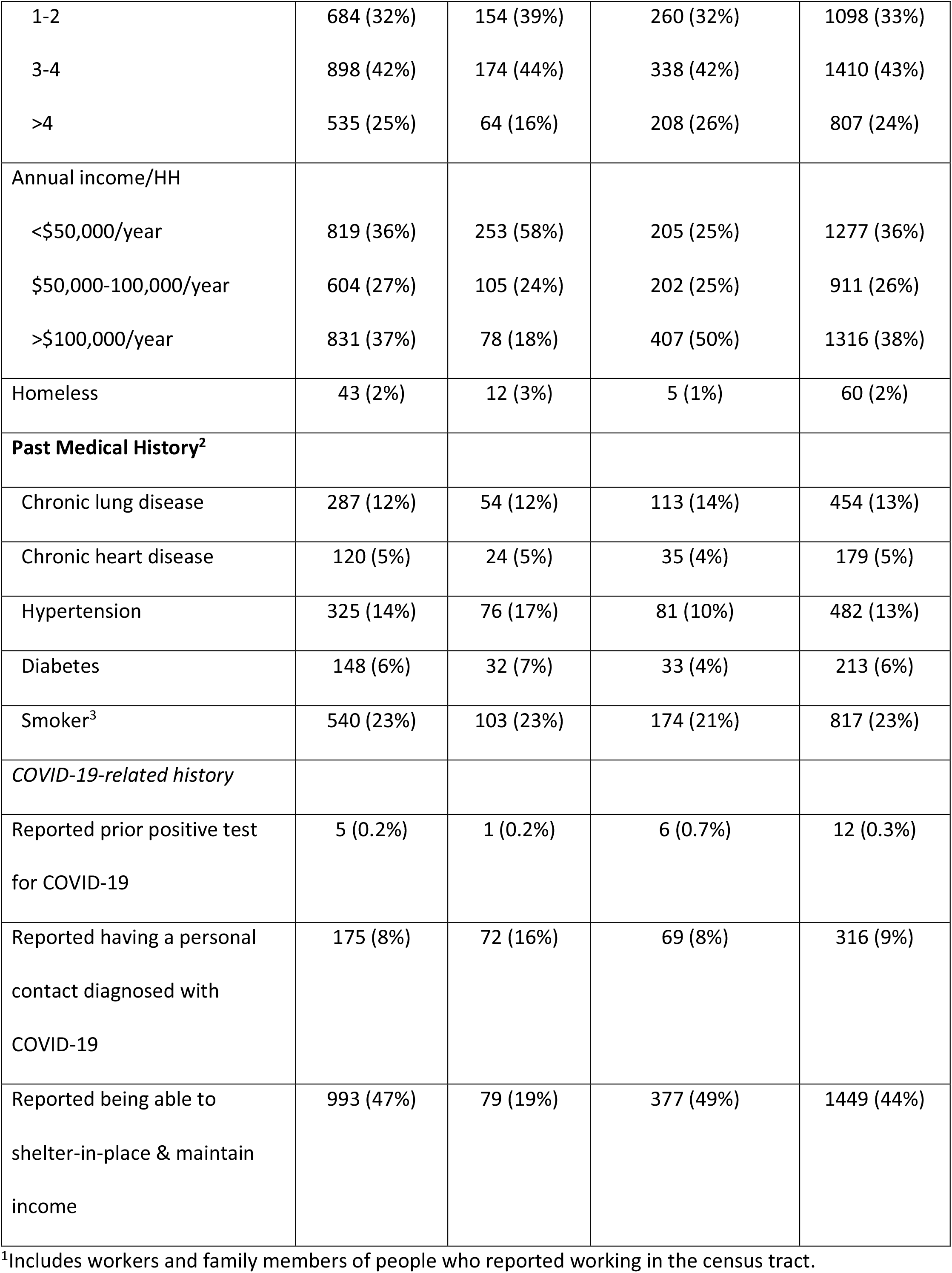
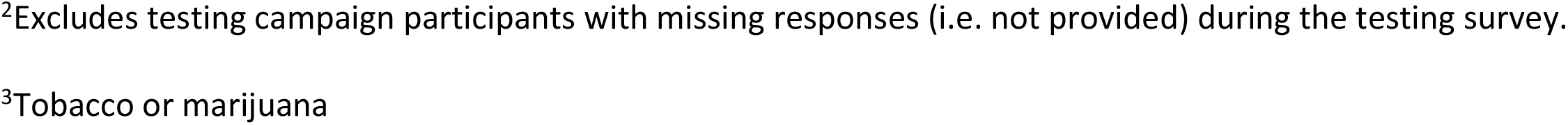
Characteristics of persons participating in a population-based SARS-CoV-2 testing campaign in the study census tract (US Census tract 022901).

### SARS-CoV-2 Infection by PCR testing

Among 3,871 tested by SARS-CoV-2 RT-PCR, 2.1% (83 people) tested PCR-positive: 1.7% (43/2,598; 95%CI: 1.2%-2.2%) of census tract residents, 6.0% (27/450; 95%CI: 4.0%-8.6%) of tract workers and 1.6% (13/823; 95%CI: 0.8%-2.7%) of residents of neighboring city blocks. Among all persons tested, 237 (6.1%) reported symptoms compatible with COVID-19 of whom 31 (13.1%) tested PCR-positive.

Among PCR-positive persons, 95% identified as Latinx, median age was 38 years (interquartile range [IQR]: 28-50 years), and 76% were male. Persons testing PCR-positive were significantly more likely than persons testing PCR-negative to identify as Latinx, report inability to shelter-in-place and maintain income, work frontline-service jobs or be unemployed, and live in households with income <$50,000/year and >3 persons/household (**Table 2**). Given that 95% of PCR-positive persons were Latinx, we limited our multivariate model to Latinx participants to evaluate risk factors for PCR-positivity within this group, and found significantly higher odds of PCR-positive infection if male (OR: 2.0, 95% CI: 1.1-3.6, p=0.02), working a frontline service job (OR: 2.6, 95% CI: 1.4-5.1, p=0.004, ref: non-frontline), household income <$50,000/year (OR: 8.9, 95% CI: 1.9-158, p=0.03, ref: >$100,000/year), or reporting a COVID-19 contact (OR: 3.6, 95%CI: 2.0-6.3, p<0.001).

**Table 2.** Characteristics of SARS-CoV-2 testing campaign participants who tested PCR-positive compared to PCR-negative, and factors associated with increased odds of PCR-positivity.

|  | PCR-positive<br>(N=83) | PCR-negative<br>(N=3,788) | Univariate risk of<br>PCR-positivity<br>(OR, 95% CI) | P value |
| --- | --- | --- | --- | --- |
| **Sex at birth (%)<sup>1</sup>** |  |  |  |  |
| Female | 20 (24%) | 1638 (47%) | *Ref* |  |
| Male | 63 (76%) | 1845 (53%) | 2.71 (1.64-4.69) | <0.001 |
| **Age category (years)** |  |  |  |  |
| 4-10 | 4 (5%) | 104 (3%) | *Ref* |  |
| 11-17 | 2 (2%) | 127 (3%) | 0.41 (0.06-2.14) | 0.3 |
| 18-50 | 60 (72%) | 2429 (64%) | 0.64 (0.26-2.15) | 0.4 |
| 51-70 | 14 (17%) | 923 (24%) | 0.39 (0.14-1.41) | 0.11 |
| >70 | 3 (4%) | 205 (5%) | 0.38 (0.07-1.76) | 0.2 |
| **Race/Ethnicity<sup>1</sup>** |  |  | *Ref: non-Hispanic/Latinx* |  |
| White/Caucasian | 1 (1%) | 1441 (42%) |  |  |
| Hispanic/Latinx | 79 (95%) | 1348 (39%) | 28.3 (11.7-93.1) | <0.001 |
| Asian/Pacific Islander | 2 (2%) | 324 (9%) |  |  |
| Other | 1 (1%) | 326 (9%) |  |  |
| **Occupation<sup>1</sup>** |  |  |  |  |
| Frontline Service Job | 47 (64%) | 902 (27%) | 6.56 (3.86-11.6) | <0.001 |
| Non-frontline Service Job | 18 (24%) | 2267 (67%) | *Ref* |  |
| Unemployed | 9 (12%) | 220 (6%) | 5.15 (2.18-11.3) | <0.001 |
| **Number of people/household<sup>1</sup>** |  |  |  |  |
| 1-2 | 10 (14%) | 1235 (37%) | *Ref* |  |
| 3-5 | 41 (58%) | 1645 (50%) | 3.08 (1.60-6.53) | 0.002 |
| >5 | 20 (28%) | 420 (13%) | 5.88 (2.79-13.2) | <0.001 |
| **Annual Household Income<sup>1</sup>** |  |  |  |  |
| >\$100,000 | 2 (3%) | 1287 (38%) | *Ref* | |
| \$50-100,000 | 7 (9%) | 889 (26%) | 5.07 (1.22-34.1) | 0.043 |
| <\$50,000 | 65 (88%) | 1182 (35%) | 35.4 (11.1-216) | <0.001 |
| **Past Medical History<sup>1</sup>** |  |  |  |  |
| Any underlying conditions | 22 (29%) | 971 (28%) | 1.02 (0.61-1.66) | 0.94 |
| **COVID-19-related history<sup>1</sup>** |  |  |  |  |
| Personal contact diagnosed with COVID-19 | 25 (32%) | 286 (8%) | 5.29 (3.19-8.57)<br>*Ref: no contacts* | <0.001 |
| Able to shelter-in-place and maintain income |  |  |  |  |
| Yes | 5 (7%) | 1417 (45%) | *Ref* | <0.001 |
| No | 64 (93%) | 1756 (55%) | 10.3 (4.58-29.6) |  |

<sup>1</sup>Excludes testing campaign participants with missing responses (i.e. not provided) during the testing survey.

Estimated point prevalence of PCR-positive infection in the census tract after adjusting for age and sex of participants in the testing campaign versus census demographics was 2.3% (95% CI: 1.2%-3.8%): 3.9% (95%CI: 2.0%-6.4%) among Latinx vs. 0.2% (95%CI: 0.0%-0.4%) among non-Latinx tract residents. Among Latinx people who worked in the census tract, unadjusted point prevalence of PCR-positive infection was 10.4% (95% CI: 7.0%-14.8%), compared to 0.0% (95% CI: 0.0%-2.0%) among non-Latinx workers.

### Clinical characteristics of PCR-positive persons

Among 83 PCR-positive persons, 43 (52%) were asymptomatic at the time of testing. Two-week follow-up was obtained for 41 participants who were asymptomatic at the time of testing: 8/41 (20%) recalled mild symptoms that had resolved by the time of testing, 10 (24%) developed symptoms after testing (pre-symptomatic) and 23 (56%) remained asymptomatic. Based on reclassified symptom status among PCR-positive people, 39/80 (49%) were symptomatic at time of testing, 8 (10%) were previously symptomatic, 10 (12.5%) were pre-symptomatic, and 23 (29%) remained asymptomatic throughout infection. One PCR-positive person (1.3%) required hospitalization.

### SARS-CoV-2 Cumulative Incidence and Recent vs. Prior Infections

Among 3,861 participants tested for SARS-CoV-2 antibodies, 3.4% (131) tested Ab-positive: 3.1% (80/2,545; 95%CI: 2.5%-3.9%) among census tract residents compared to 7.7% (34/442; 95%CI: 5.4%-10.6%) among tract workers and 2.1% (17/829; 95%CI: 1.2%-3.3%) among adjacent city block residents. Estimated cumulative incidence (Ab or PCR-positive) among tract residents was 4.4% (95% CI: 3.2%-5.6%) after adjusting for test characteristics and 6.1% (95%CI: 4.0%-8.6%) after further adjusting for participation (**Table S1**).

Among all infections detected by PCR or Ab, 26% (48/182) were recent infection. 53% (96/182) were prior infection. Of the remaining infections, 18% (32/182) were PCR-positive/Ab-positive, and 3% (6/182) had PCR or antibody testing alone (**Table S2**). Compared to individuals with prior infection, people with recent infection were significantly more likely to be of Latinx ethnicity, report inability to shelter-in-place and maintain income, work frontline service jobs or be unemployed, and live in households with income <$50,000/year (**Table 3**). Moreover, 47% vs. 89% of prior vs. recent infections, respectively, occurred in households with income <$50,000/year (**Table S2**).

**Table 3.**
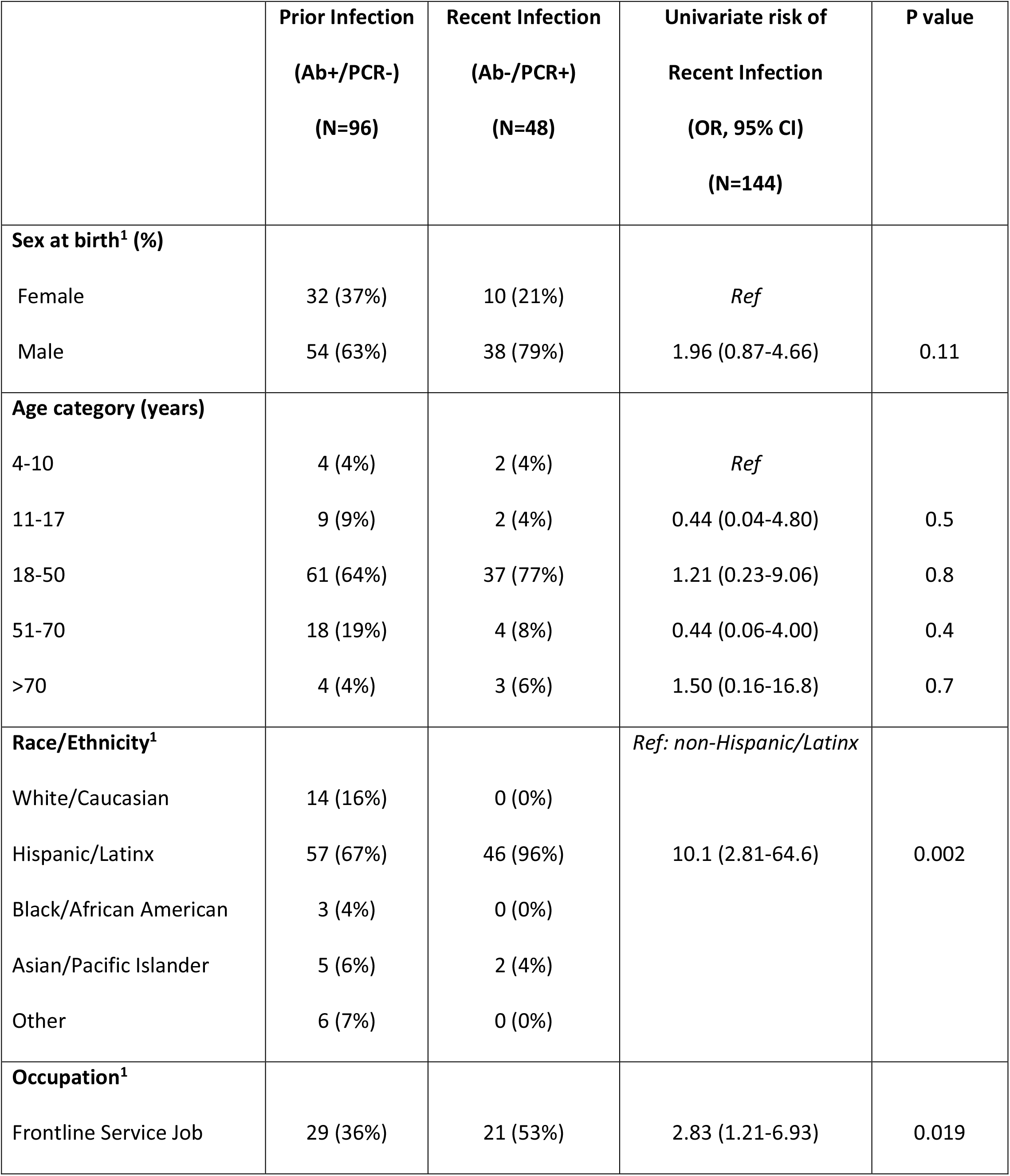
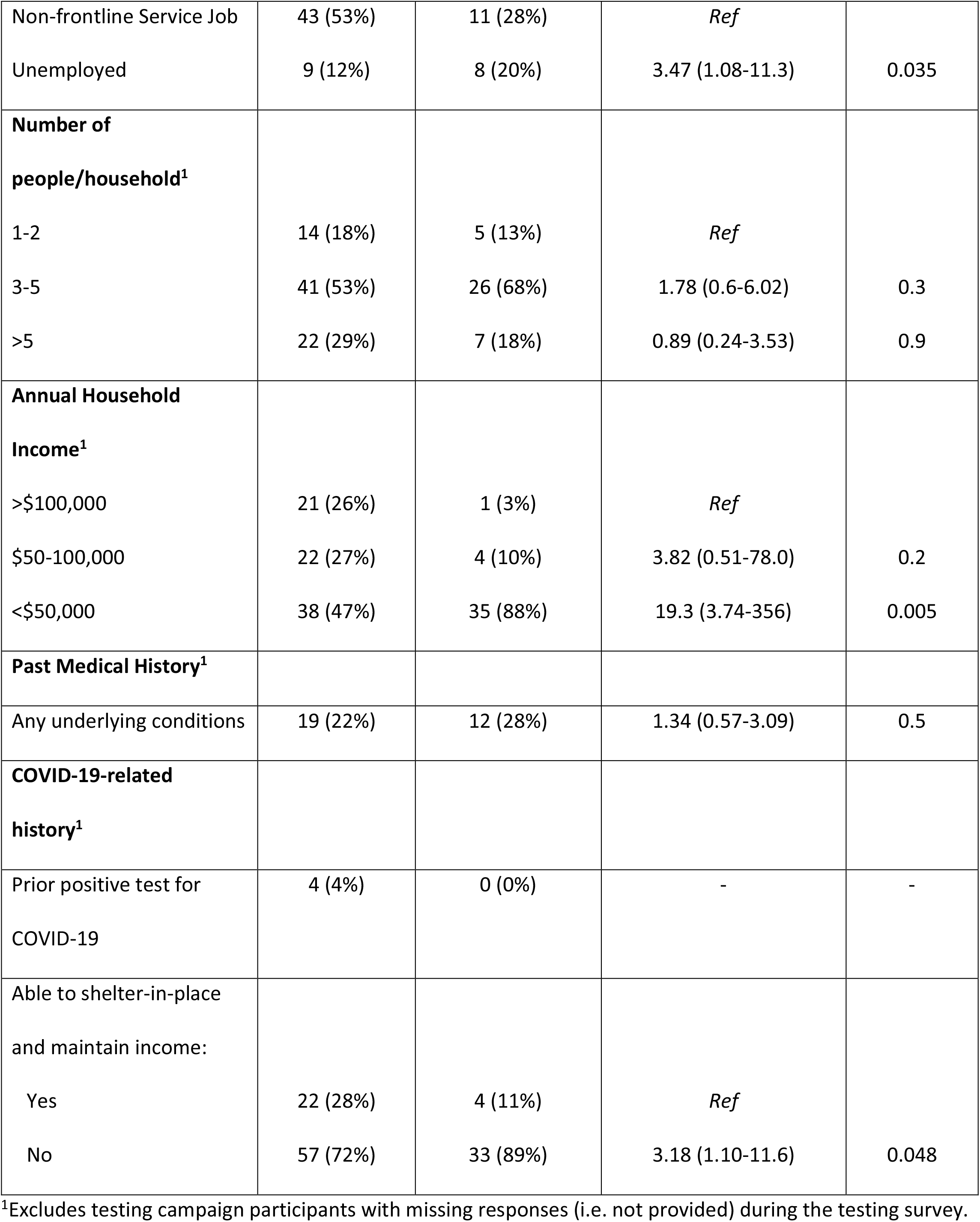
Comparison of prior (Ab+/PCR-) vs. recent infection (Ab-/PCR+) infection among persons participating in a population-based SARS-CoV-2 testing campaign.

### SARS-CoV-2 RNA levels and phylogenetic analysis

Median levels of virus as estimated by RT-PCR cycle thresholds were significantly higher among PCR-positive/Ab-negative persons compared to PCR-positive/antibody-positive persons, supporting our classification of recent infection (**Figure 1**). Among recently-infected individuals, median levels of virus by RT-PCR cycle threshold did not differ significantly between symptomatic (24, IQR: 19-25, range 11-35; N=27) and asymptomatic (24, IQR: 19-26, range 16-32; N=10; p=0.98) persons (**Figure 1**); additional comparisons by subgroup are in **Figure S1**.

**Figure 1:**
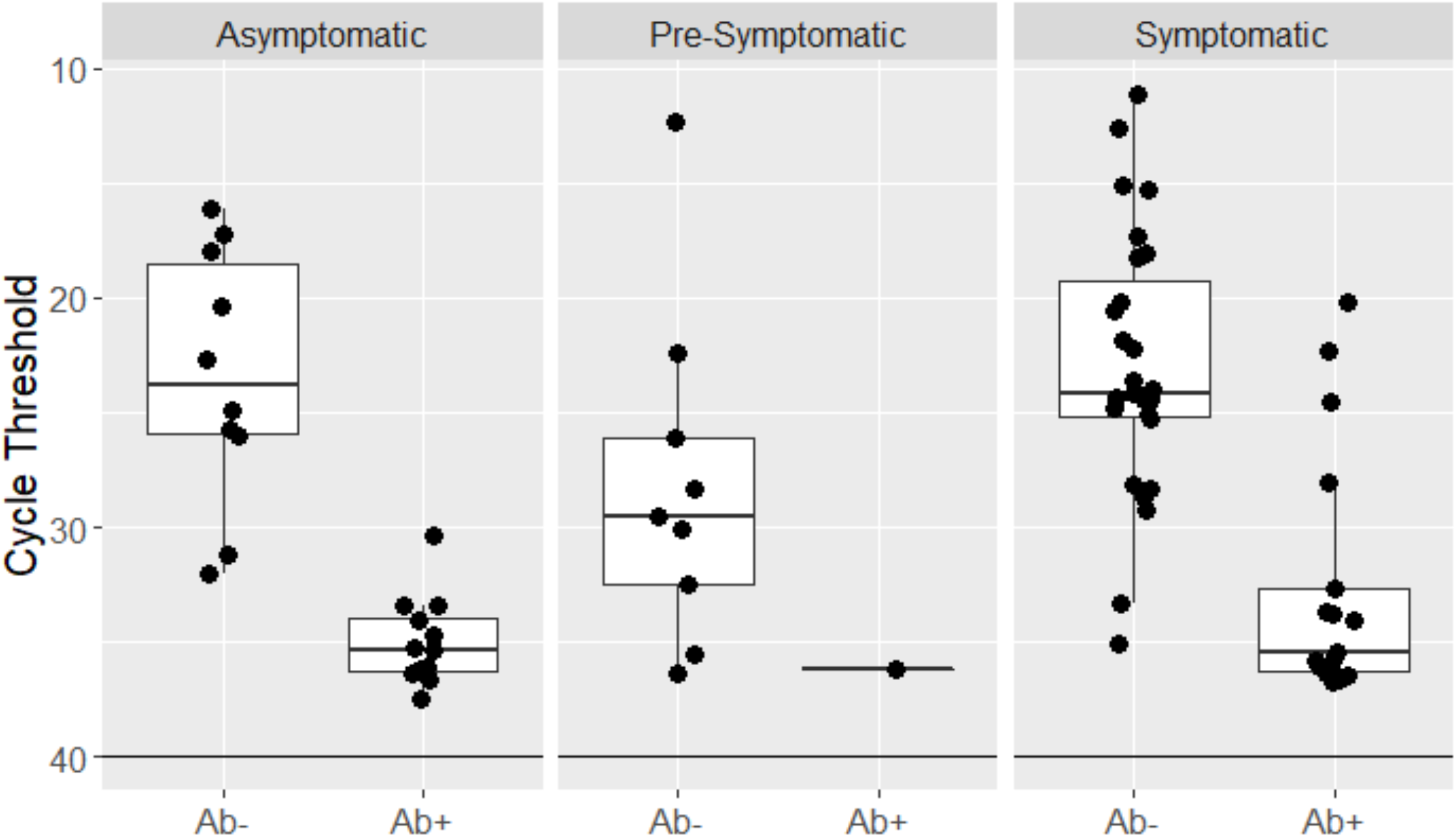
Quantitative levels of virus among participants with PCR-positive SARS-CoV-2 infections (N=80) by classification as asymptomatic, pre-symptomatic, and symptomatic for COVID-19 disease as determined over longitudinal follow-up (2 weeks post-testing), and stratified by antibody status with PCR+/Abpersons considered consistent with recent infection.

We recovered SARS-CoV-2 genomes from 59% (49/83) of the PCR-positive RNA samples. The recovered genomes were diverse and phylogenetically intermixed with samples from across San Francisco, including representatives from five globally circulating clades, showing multiple independent introductions (**Figure 2, Panel A**). Overall, 58% of PCR-positive participants shared a home with another PCR-positive participant identified in the testing campaign: sequences from such households were consistent with within-household transmission (**Figure 2, Panel B)**, with no variants detected in 65% of household links (N = 11/17, 95% CI 41%-83%). We found no significant population structure separating the Mission district samples from the rest of San Francisco (p=0.19).

**Figure 2:**
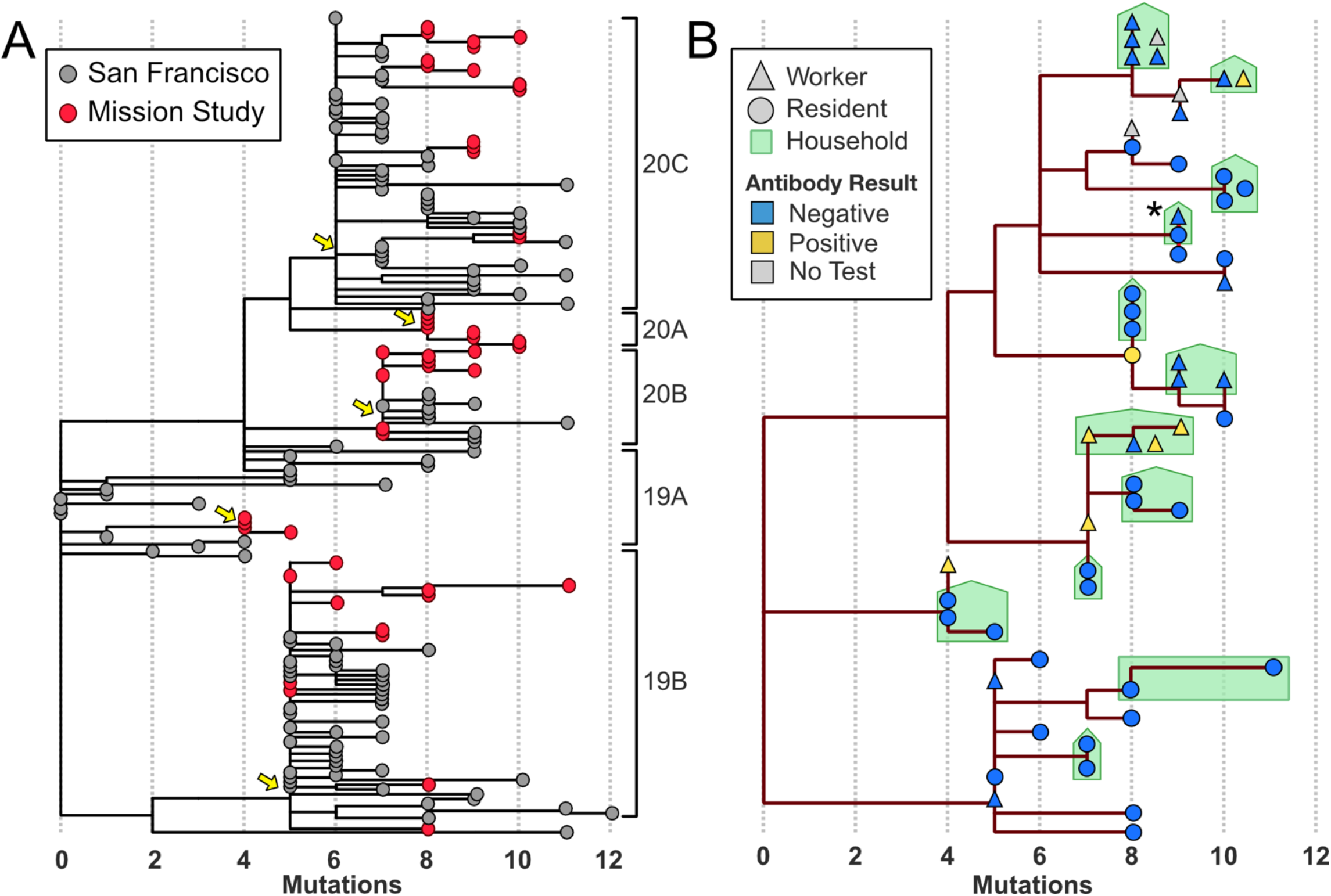
Viral genomic diversity among PCR-positive participants. Panel A: phylogenetic tree containing Mission district samples (red) and other San Francisco samples (grey), x-axis marks the number of mutations with respect to the reference genome from Wuhan. Yellow arrows mark introductions of five major global clades (right brackets) to the study population. Panel B: tree subset to Mission district samples. Shape indicates district resident or worker and color indicates antibody status. Households with multiple PCR-positive persons are drawn in green and include markers for samples from which genomes could not be recovered. Asterisk marks a household outside of the district in which an unhoused person in the district spent time.

## Discussion

We found stark ethnic and economic disparities in who is at risk for ongoing infection six-weeks into San Francisco’s shelter-in-place ordinance. The estimated point prevalence of PCR-positive infection among Latinx residents (3.9%) was twenty times that of non-Latinx residents (0.2%). Perhaps even more striking was that recent infections were concentrated almost exclusively among low-income, Latinx people working frontline jobs, whereas prior infections occurred among more ethnically and economically diverse individuals. In addition, the majority of PCR-positive infections were asymptomatic at the time of testing, and recently infected individuals had high levels of virus regardless of symptoms. These data show that San Francisco’s COVID-19 epidemic has continuing transmission in subgroups of the city population that require urgent attention.

Heterogeneity among populations most affected by the COVID-19 epidemic exists across the U.S., within states and cities, and even within neighborhoods as shown here. Population-level epidemiologic data coupled with phylogenetic analyses can help identify, track and inform testing strategies, public health policies and measures to mitigate health and economic effects. Low-barrier, community-mobilized testing is foundational to these efforts. We sought to overcome testing barriers in this census tract through our partnership with the community-led Latino Task Force in San Francisco, who provided trusted explanations about COVID-19 and messaging on the importance of testing to the community. Through this approach, we were able to test a large proportion of the population in a short period of time.

We determined that during shelter-in-place, COVID-19 transmission became increasingly concentrated in Latinx community members. The risk factors driving recent transmission among Latinx residents were largely economic and highly correlated: low-income residents working frontline jobs who could not shelter-in-place and maintain their income. Transmission was amplified in Latinx multi-generational and multi-family households – a byproduct of skyrocketing rental costs in the city. These economic drivers and ethnic disparities observed at the community level here are reflected in COVID-19 hospitalizations and deaths widely reported in the US.^10,20^

We observed high sequence diversity of SARS-CoV-2 infections in the census tract, similar to the diversity seen in San Francisco more broadly, suggestive of multiple independent introductions over time. Our data suggest that most recent infections during shelter-in-place are due to acquisition of virus when working or seeking work in the census tract, with subsequent within-home transmission in high-density, low-income households. These findings should help dispel common, dangerous pitfalls in interpreting ethnic disparities in infection, such as biological explanations, supposed community behaviors or stigmatizing communities as transmission “hot spots” about which others have cautioned.^11^

Our results also highlight the importance of SARS-CoV-2 testing in both symptomatic and asymptomatic individuals. Symptom-based testing would have failed to detect over 40% of PCR-positive infections in this community, many of whom had high levels of virus. Overall, 29% of PCR-positive infections never developed symptoms, slightly lower than the proportion found (43%) in a population-based SARS-CoV-2 screening study from Iceland.^21^ In our study, recently infected people had high levels of virus that did not differ significantly based on symptoms. In addition, only one infected individual required hospitalization, suggesting the vast majority would not have been diagnosed without community-based testing. The clear implication of these findings is that testing strategies limited to symptomatic individuals and those seeking testing at health centers alone, will fail to limit transmission.

These results have several implications moving forward as shelter-in-place restrictions are lifted. First, more efforts are needed to address uncontrolled epidemics among sub-populations, especially vulnerable populations such as the Latinx community highlighted here. Expanded targeted, but community-led and mobilized, low-barrier testing not based on symptoms is needed. Testing need to be coupled with social protection of job security and economic support for self-isolation and quarantine (i.e. “test and respond”) and culturally responsive contact tracing. Our testing campaign contributed to policy change in San Francisco, with the mayor announcing on May 4, 2020 that essential workers would be eligible for free SARS-CoV-2 testing regardless of symptoms,^22^ and then on May 28, 2020 that low-wage workers with COVID-19 would be provided funds to stay home and isolate (“Right to Recover”).^23^ In parallel, longitudinal, population-based cohorts that couple epidemiologic data with PCR and antibody testing and viral sequencing can provide evidence of effectiveness of public health measures and viral introductions over time, enabling evidence-based responses in a dynamic landscape.

Our study has several limitations. SARS-CoV-2 PCR tests do not detect all cases and antibody sensitivity may be lower in asymptomatic infection which could have resulted in underestimation of cumulative incidence. False positive antibody results could result in overestimation of cumulative incidence and misclassification of prior infections. Fingerstick sampling could also impact antibody test performance. However, the EUA antibody test we used has been shown to have a high sensitivity (96.9-100%) at ≥17-22 days post symptom onset, and high specificity (≥99.6%) with venous drawn plasma,^24,25^ and our estimates of cumulative incidence accounted for sensitivity and specificity of the PCR and antibody assays used. Second, selection bias in who chose to test may have affected our estimates. Although we adjusted for demographic differences between participants and community composition based on 2018 American Community Survey data, these data may not fully reflect tract demographics in 2020. However, population-based testing in a census tract allowed for greater clarity in understanding who did not participate. Lastly, we relied on self-reported symptoms and survey responses, which may have resulted in misclassification. With follow-up of PCR-positive participants over two weeks, we were able to further explore symptom status, allowing for monitoring and reclassification.

In conclusion, improving access to SARS-CoV-2 testing, regardless of symptoms, through community-led, low-barrier testing programs in vulnerable communities, coupled with economic support and protections for low-income workers during isolation and quarantine are urgently needed to reduce community transmission and address the massive disparities in SARS-CoV-2 infection observed in the U.S.

## Data Availability

The data that support the findings will be available following publication.

## Acknowledgements

We thank the community members from the census tract studied for their generous participation in our study during San Francisco’s shelter-in-place ordinance. We gratefully acknowledge the contributions of all of the study volunteers who made community-based testing possible, and all the members of the Latino Task Force for COVID-19 who led community mobilization efforts, community outreach and education and dissemination of our findings to the community. We thank Chesa Cox, MPH for logistical and administrative support, Stacie Powers for her generous donation of the Brava Theater space to operate as headquarters for the study volunteers and the Community Wellness Team. We thank San Francisco Recreation and Parks and the San Francisco Unified School District for generously allowing us to offer community-based testing at two public parks and two public schools. We thank Supervisor Hillary Ronen, the San Francisco Mayor’s Office, Dr. Grant Colfax Director of San Francisco Department of Public Health, Elaine Forbes, Executive Director, Port of San Francisco as well as Naomi Kelly, SF City Administrator, Dr. Susan Ehrlich CEO of Zuckerberg San Francisco General Hospital and the Ward 86 Clinic staff for their efforts in support of this community-based testing project. We thank Andrew Kobylinski and Primary tech for assistance in their virtual support platform and online results reporting. We also wish to thank Ana Vallari, Ana Olivo, Barb Harris, and Chris Lark from Abbott Laboratories for helping conduct the antibody assays for this study.

## Supplementary Appendix

Supplement to manuscript: SARS-CoV-2 Community Transmission During Shelter-in-Place in San Francisco

### Supplementary Methods

Statistical approach to point estimates and 95% Confidence intervals for cumulative incidence, adjusted for test characteristics and sample participation

#### Overview of Approach

We take as our target of inference the cumulative probability of infection by SAR-COV-2 up to three days prior to the testing period.

Cumulative incidence unadjusted for test characteristics is estimated as the empirical proportion of participants with either a positive antibody test, a positive PCR test, or both. Corresponding unadjusted 95% CIs are estimated using an exact binomial approach. In this supplement, we describe the approach used to adjust both the point estimate and 95% CI to account for imperfect test sensitivity and specificity, as well as uncertainty in the estimates of these test characteristics. We further describe how, among census tract residents, additional adjustment for demographic differences between 2018 census tract composition (Manson, 2019) and study participation is incorporated. Uncertainty due to both forms of adjustment is estimated based on a bootstrap-based approach, related to the methods described in Bendavid, et al (2020), Cherain (2020), and Gelman (2020), and described in further detail below.

Use of the bootstrap approach requires estimates of test sensitivity and specificity, together with measures of the uncertainty in those estimates. Because our estimate of cumulative incidence is based on a composite outcome of two assays (PCR and antibody), we thus require an estimate of the specificity and sensitivity of the composite test and corresponding uncertainty estimates.

#### Estimation of specificity (and uncertainty of estimate) for composite cumulative incidence outcome

Specificity of the composite testing outcome (Antibody-positive or PCR-positive) can be estimated based on estimates of each of these tests in turn. For PCR specificity, we use data from a recent study in Bolinas in which 1845/1845 persons tested with the identical PCR assays under identical conditions tested negative. Determining specificity requires knowledge of the true number of uninfected persons, which is unknown but very unlikely to be less than 1827^1^, PCR point estimate specificity is estimated using a conservative approach applied to these data: 0.5^1/1827^ = 99.96% (Marill, 2017). For Antibody specificity, we use test characteristics provided by the manufacturer in the FDA package insert, in which 1066/1070 persons tested negative, implying a specificity point estimate of 99.63% (95% CI: 99.05%, 99.90%).

We assume that for an uninfected person, the probability of a negative PCR test is independent of the probability of a negative Antibody test, so joint specificity = PCR specificity * Antibody specificity. Together, these data provide a point estimate of the specificity of testing Antibody-positive and/or PCR-positive of 99.59% (95% CI 99.16%, 99.91%). The confidence interval is generated by a bootstrap procedure:

1. Repeat 5000 times:
  a. Create a vector of 1827 Bernoulli variables with probability of correct 0.5^1/1827^. Take a bootstrap sample of size 1827 with replacement (Marill, 2017)
  b. Take an independent bootstrap sample of 1066 correct and 4 incorrect.
  c. The joint result is correct if both bootstrap samples are correct.
2. Find the 2.5^th^ and 97.5^th^ quantile of the joint result.

#### Estimation of sensitivity (and uncertainty of estimate) for composite cumulative incidence outcome

Estimation of the sensitivity of the composite test (i.e., probability of testing Antibody-positive and/or PCR-positive given an infection up to three days before testing) is considerably more complex for several reasons.

- First, sensitivity varies profoundly for both assays as a function of time since infection. Based on current best available data, PCR sensitivity likely peaks around Day 8 after infection (∼Day 3 after symptom onset), and declines quickly; however, it remains greater than 0 for at least three weeks after infection (Kucirka, et al, 2020). Antibody sensitivity may be zero (0 positive, 4 negative) during the first two days of symptoms, then increases to close to perfect sensitivity (88 positive, 0 negative) after day 14 following symptom onset (per FDA package insert). In estimates below, we use these sources for estimated time-varying assay-specific sensitivity. We further assume a sensitivity of zero in the initial three days following infection. Any individuals in our sample who were infected within the three days prior to testing and tested PCR-positive on PCR will thus contribute to an overestimate of cumulative incidence; however, the impact of such events, if any, is expected to be very small.
- Second, because assay sensitivity varies by time, the sensitivity of the combined composite assay on any given calendar date depends on the distribution of time since infection on that date among persons who have been infected with SARS-COV-2 in the underlying population. This distribution in turn depends on the epidemic course to date in the underlying population. For example, high early growth rates followed by lower recent growth rates will increase the proportion of true positives with longstanding infection; recent high growth rates will increase the proportion of true positives who have been recently infected. The true historical underlying growth rate in this population is unknown. Here, we assume a set of time-varying growth rates generally consistent with regional hospitalization curves, and perform sensitivity analysis under variations in these assumed rates.
- Third, available data on time dependent-sensitivity curves have, to date, been drawn almost exclusively from symptomatic individuals. It is possible that sensitivity for either or both assays may be lower among asymptomatic persons. In the absence of data, we assume equivalent time-specific sensitivity among symptomatic and asymptomatic persons for each assay. The approach presented can be readily adopted (and we provide code to do so) if additional data on differential sensitivity become available.

With these basic assumptions, we implement the following approach to estimating the sensitivity of the joint testing outcome, together with uncertainty.

Repeat 5000 times

1. Calculate Antibody sensitivity:
  a. For Antibody sensitivity by day since symptoms, sample with replacement using data from the FDA insert for days 1-13 of symptoms: (0/4 positive antibody tests on days 1-2 of symptoms; 2/8 positive antibody tests on days 3-7 of symptoms; 19/22 positive antibody tests on days 8-13 of symptoms).
  b. For day ≥14 of symptoms, the FDA insert reports 88/88 positive antibody tests^2^. Create a vector of 88 Bernoulli variables with probability of correct 0.5^1/88^. Take a bootstrap sample of size 88 with replacement (Marill, 2017)
  c. Antibody sensitivity by day since symptoms is estimated as the mean of the bootstrap sample
2. Calculate PCR sensitivity
  a. For PCR sensitivity by day since infection, we use the results of Kucirka et al (2020), posted on https://github.com/HopkinsIDD/covidRTPCR. Sample one of the 5000 posterior draws of these results. Kucirka, et al provide estimates of PCR sensitivity from day 1 to 21 since infection.
  b. We assume sensitivity on day 31 is zero and use linear interpolation to estimate PCR sensitivity on days 22 to 30.
3. Assume that infections start on Feb 20, grow 23% per day (R0 ∼ 3.5), and that the growth rate than decreases linearly starting March 7 to 5% per day on March 23 (Re ∼ 1.2), reflecting the imposition of a Shelter In Place ordinance in San Francisco. These reproductive numbers are supported by fitting an SEIR model to San Francisco hospitalization data. Calculate the fraction of infections by day under these growth rates. Assume that symptoms start on 5^th^ day of infection (as in Kucirka, et al).
4. Assume that for a truly infected person, the probability of a positive PCR test is independent of the probability of a positive Antibody test. Then joint sensitivity by day since infection = PCR sensitivity + Antibody sensitivity - PCR sensitivity * Antibody sensitivity
5. As noted above, we take as our target of inference cumulative probability of infection by SAR-COV-2 up to three days prior to the testing period, and thus assume sensitivity of zero in the initial three days following infection. Kucirka, et al estimate sensitivity of less than 0.01% on day 1, 0.4% on day 2, 6.6% on day 3, but confidence intervals include zero on all three days.
6. Joint sensitivity integrated over days since infection is calculated by summing the product of fraction of infections by day and joint sensitivity by day.

The 5000 repetitions give a mean joint sensitivity of the combined Antibody and PCR assays of 88.4% with a standard deviation of 2.4%. Only this final mean and standard deviation are used in all adjusted estimates in main text of the paper. Because there were many assumptions in the estimation of these two quantities, we conducted sensitivity analysis considering a range of point and variance estimates for composite test sensitivity; results are shown in Table.

#### Bootstrap-based approach to adjust point estimate and confidence interval

Given the above inputs – estimated sensitivity and corresponding 95% CI of the joint test, and specificity estimates of the PCR and Antibody Tests with corresponding numbers of tests used to generate these estimates – we implemented the following bootstrap-based procedure to adjust the point estimates and generate confidence intervals accounting for uncertainty in test characteristics (and optionally allowing for adjustment for participation).

For each bootstrap iteration *j* in 1 to 5000

1. Draw composite test sensitivity, *sens*^*j*^, from a normal distribution with mean 88.4% and standard deviation of 2.4%
2. Draw composite test specificity:
  a. Create a vector of 1827 Bernoulli variables with probability of correct 0.5^1/1827^. PCR specificity = mean of a bootstrap sample of size 1827 with replacement. (Marill, 2017)
  b. Antibody specificity = mean of a bootstrap sample of 1066 correct and 4 incorrect.
  c. Composite test specificity = *spec*^*j*^ = PCR specificity * Antibody specificity
3. Draw *H* household at random (with replacement), and add all members of that household to the bootstrap sample, where *H* is the total number of households. Define *qgj* as the proportion of people with positive test results in the *j*^th^ iteration in the race-sex-age cell *g*.
4. For each race-sex-age cell *g*, convert test results to expected rate of actual prevalence using the following formula (Bendavid et al, 2020): *pgj* = max(0, (*qgj* + *spec*^*j*^ - 1) / (*sens*^*j*^ + *spec*^*j*^ - 1))
5. Calculate the expected prevalence by summing over all of the groups, weighting group *g* by the group’s census population (*Ng*) divided by the total census population (*N*).

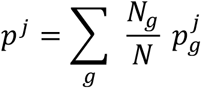

The 95 percent confidence interval is defined by the 2.5^th^ and 97.5^th^ percentiles of the observed values of *p*.

Table. Sensitivity analysis, showing adjusted point estimates and 95% CIs under varying assumptions on time-dependent sensitivity of the time-dependent Antibody and PCR tests. (Primary estimated, reported in main text, shown in blue.)

| Composite Sensitivity |  | Infection among Residents, adjusted for demographics and test characteristics |  |  |
| --- | --- | --- | --- | --- |
| Mean | Std Dev | Point Est | CI_low | CI_high |
| 70.0% | 1.0% | 7.7% | 5.0% | 10.8% |
| 70.0% | 2.4% | 7.7% | 5.0% | 10.9% |
| 70.0% | 5.0% | 7.7% | 5.0% | 11.1% |
| 80.0% | 1.0% | 6.7% | 4.4% | 9.5% |
| 80.0% | 2.4% | 6.7% | 4.4% | 9.5% |
| 80.0% | 5.0% | 6.7% | 4.3% | 9.7% |
| 88.4% | 1.0% | 6.1% | 4.0% | 8.6% |
| 88.4% | 2.4% | 6.1% | 4.0% | 8.6% |
| 88.4% | 5.0% | 6.1% | 3.9% | 8.7% |
| 95.0% | 1.0% | 5.7% | 3.7% | 8.0% |
| 95.0% | 2.4% | 5.7% | 3.7% | 8.0% |
| 95.0% | 5.0% | 5.7% | 3.7% | 8.1% |

## Supplementary Methods

Bioinformatics and Genomic Analyses

Consensus genomes were generated by first using minimap2^1^ to align raw reads to the reference genome MN908947.3, then using samtools,^2^ mpileup and ivar^3^ to generate consensus genomes. We used the augur/auspice^4^ toolkit with mafft,^5^ iqtree,^6^ and treetime^7^ to construct and visualize maximum likelihood phylogenies. The inferred topology had a lower bound of 9 transmission events into and out of the Mission, based on a min-cut separating the Mission district from other San Francisco samples. Branches on the topology with zero inferred mutations were collapsed into polytomies for the min-cut analysis. To test for population structure, we used the Python package scikit-allel to compute Hudson’s F _ST_^8^ between the Mission District study census tract and sequences from GISAID^9^ representing the rest of San Francisco (Supplemental Table S3), including at most 1 sample per household (the first lexicographically), and including all variant sites in the multiple alignment with at most 5 missing alleles. We failed to reject the null hypothesis of no population structure, computing FST=.007 with a p-value of 0.192 based on a permutation test with 10,000 random rearrangements of the sample labels.

---

**Table S1:** Summary of estimated point prevalence of PCR-positive SARS-CoV-2 infection and estimated cumulative incidence of SARS-CoV-2 infection among census tract residents, persons who work but do not live in the census tract, and residents from city blocks adjacent to the study census tract.

|  | **Census Tract Residents** | **Workers** | **Adjacent City Blocks** | **All** |
| --- | --- | --- | --- | --- |
| **PCR-positive (unadjusted)** | 43/2,598<br>(1.7%, 95%CI: 1.2%-2.2%) | 27/450<br>(6.0%, 95%CI: 4.0%-8.6%) | 13/823<br>(1.6%, 95%CI: 0.8%-2.7%) | 83/3,871<br>(2.1%, 95%CI: 1.7%-2.7%) |
| **Estimated point prevalence of PCR-positive in the census tract<sup>1</sup>** | 2.3% (95% CI: 1.2-3.8%) | N/A | N/A | N/A |
| **Antibody-positive (unadjusted)** | 80/2,545<br>(3.1%, 95%CI: 2.5%-3.9%) | 34/442<br>(7.7%, 95%CI: 5.4%-10.6%) | 17/829<br>(2.1%, 95%CI: 1.2%-3.3%) | 131/3816<br>(3.4%, 95%CI: 2.9%-4.1%) |
| **PCR-positive or Antibody-positive** | 106/2491<br>(4.3%, 95%CI: 3.5%-5.1%) | 51/437<br>(11.7%, 95%CI: 8.8%-15.1%) | 25/812<br>(3.1%, 95%CI: 2.0%-4.5%) | 182/3740<br>(4.9%, 95%CI: 4.2%-5.6%) |
| **Estimated cumulative incidence of infection (PCR or Ab-positive)<sup>2</sup>** | 106/2491<br>(4.4%, 95%CI: 3.2%-5.6%) | 51/437<br>(12.8%, 95%CI: 8.4%-17.6%) | 25/812<br>(3%, 95%CI: 1.5%-4.7%) | 182/3,740<br>(5.1%, 95%CI: 4.0%-6.2%) |
| **Estimated cumulative incidence of infection (PCR or Ab-positive) in the census tract<sup>3</sup>** | 6.1% (95%CI: 4.0%-8.6%) | N/A | N/A | N/A |

<sup>1</sup>Adjusted for age, sex and race/ethnicity of testing participants vs. census demographics (excludes 339 participants tested by PCR who were missing age, sex, or race/ethnicity)
<sup>2</sup>Adjusted for test characteristics
<sup>3</sup>Adjusted for test characteristics and age, sex, and race/ethnicity of testing participants vs. census demographics (excludes 319 participants tested by antibody who were missing age, sex, or race/ethnicity)
N/A: Not applicable

**Table S2.** Characteristics of participants with SARS-CoV-2 infection diagnosed by PCR or Antibody (N=144 total with classification as recent vs. prior infection), and antibody positivity by participant characteristics.

|  | **Prior Infection**<br>**(Ab+/PCR-)** | **Prior or Recent Infection**<br>**(Ab+/PCR+)** | **Recent Infection**<br>**(Ab-/PCR+)** | **Antibody Positive among tested, row%** |
| --- | --- | --- | --- | --- |
| **N** | 96 | 32 | 48 | 131/3816 (3.4%) |
| **Sex at birth<sup>1</sup> (%)** |  |  |  |  |
| Female | 32 (37%) | 9 (28%) | 10 (21%) | 42/1643 (2.6%) |
| Male | 54 (63%) | 23 (72%) | 38 (79%) | 79/1871 (4.2%) |
| **Age category, in years** |  |  |  |  |
| 4-10 | 4 (4%) | 2 (6%) | 2 (4%) | 6/109 (5.5%) |
| 11-17 | 9 (9%) | 0 (0%) | 2 (4%) | 9/135 (6.7%) |
| 18-50 | 61 (64%) | 21 (66%) | 37 (77%) | 83/2466 (3.4%) |
| 51-70 | 18 (19%) | 9 (28%) | 4 (8%) | 29/916 (3.2%) |
| >70 | 4 (4%) | 0 (0%) | 3 (6%) | 4/190 (2.1%) |
| **Race/Ethnicity** |  |  |  |  |
| White/Caucasian | 14 (16%) | 1 (3%) | 0 (0%) | 16/1433 (1.1%) |
| Hispanic/Latinx | 57 (67%) | 30 (94%) | 46 (96%) | 89/1389 (6.4%) |
| Black/African American | 3 (4%) | 0 (0%) | 0 (0%) | 3/79 (3.8%) |
| Asian/Pacific Islander | 5 (6%) | 0 (0%) | 2 (4%) | 5/325 (1.5%) |
| Other | 6 (7%) | 1(3%) | 0 (0%) | 7/244 (2.9%) |
| **Occupation** |  |  |  |  |
| Frontline job | 29 (36%) | 23 (74%) | 21 (53%) | 52/924 (5.6%) |
| Non-frontline job | 43 (53%) | 7 (23%) | 11 (28%) | 53/2258 (2.3%) |
| Unemployed | 9 (12%) | 1 (3%) | 8 (20%) | 10/230 (4.3%) |
| **Number people/household (HH)** |  |  |  |  |
| 1-2 | 14 (18%) | 4 (13%) | 5 (13%) | 19/1230 (1.5%) |
| 3-5 | 41 (53%) | 14 (47%) | 26 (68%) | 56/1656 (3.4%) |
| >5 | 22 (29%) | 12 (40%) | 7 (18%) | 35/432 (8.1%) |
| **Annual HH Income** |  |  |  |  |
| >\$100,000 | 21 (26%) | 1 (3%) | 1 (3%) | 23/1281 (1.8%) |
| \$50-100,000 | 22 (27%) | 3 (10%) | 4 (10%) | 25/889 (2.8%) |
| <\$50,000 | 38 (47%) | 27 (87%) | 35 (88%) | 67/1225 (5.5%) |
| **Hispanic/Latinx Annual HH Income** |  |  |  |  |
| >\$100,000 | 7 (13%) | 0 (0%) | 1 (3%) | 7/155 (4.5%) |
| \$50-100,000 | 12 (23%) | 2 (7%) | 3 (8%) | 14/334 (4.2%) |
| <\$50,000 | 34 (64%) | 27 (93%) | 34 (89%) | 63/808 (7.8%) |
| **White/Caucasian Annual HH Income** |  |  |  |  |
| >\$100,000 | 6 (46%) | 1 (100%) | 0 (0%) | 8/808 (1.0%) |
| \$50-100,000 | 7 (54%) | 0 (0%) | 0 (0%) | 7/373 (1.9%) |
| <\$50,000 | 0 (0%) | 0 (0%) | 0 (0%) | 0/209 (0%) |
| **Past Medical History** |  |  |  |  |
| Any underlying conditions | 19 (22%) | 9 (29%) | 12 (28%) | 30/968 (3.1%) |
| **COVID-19-related history** |  |  |  |  |
| Prior positive test for COVID-19 | 4 (4%) | 5 (16%) | 0 (0%) | 9/11 (81.8%) |
| Able to shelter-in-place and maintain income? |  |  |  |  |
| Yes | 22 (28%) | 1 (4%) | 4 (11%) | 23/1411 (1.6%) |
| No | 57 (72%) | 28 (97%) | 33 (89%) | 88/1786 (4.9%) |

**Table S3.** GISAID sequences used for phylogenetic analysis. We gratefully acknowledge the following Authors from the Originating laboratories responsible for obtaining the specimens and the Submitting laboratories where genetic sequence data were generated and shared via the GISAID Initiative, on which this research is based. All submitters of data may be contacted directly via www.gisaid.org.

| Name | Accession | Date | Originating laboratory | Submitting laboratory | Authors |
| --- | --- | --- | --- | --- | --- |
| USA/CA-CZB043/2020 | EPI_ISL_429048 | 3/25/2020 | UCSF Clinical Microbiology Laboratory | Chan-Zuckerberg Biohub | CZB Cliahub Consortium et al |
| USA/CA-CZB0721/2020 | EPI_ISL_429020 | 3/31/2020 | UCSF Clinical Microbiology Laboratory | Chan-Zuckerberg Biohub | CZB Cliahub Consortium et al |
| USA/CA-CZB-1100/2020 | EPI_ISL_444058 | 4/28/2020 | UCSF Clinical Microbiology Laboratory | Chan-Zuckerberg Biohub | CZB Cliahub Consortium et al |
| USA/CA-CZB0151/2020 | EPI_ISL_429061 | 4/8/2020 | UCSF Clinical Microbiology Laboratory | Chan-Zuckerberg Biohub | CZB Cliahub Consortium et al |
| USA/CA-CZB011b/2020 | EPI_ISL_429063 | 3/25/2020 | UCSF Clinical Microbiology Laboratory | Chan-Zuckerberg Biohub | CZB Cliahub Consortium et al |
| USA/CA-CZB099/2020 | EPI_ISL_429071 | 3/28/2020 | UCSF Clinical Microbiology Laboratory | Chan-Zuckerberg Biohub | CZB Cliahub Consortium et al |
| USA/CA-CZB076/2020 | EPI_ISL_429057 | 3/28/2020 | UCSF Clinical Microbiology Laboratory | Chan-Zuckerberg Biohub | CZB Cliahub Consortium et al |
| USA/CA-CZB0154/2020 | EPI_ISL_429047 | 4/8/2020 | UCSF Clinical Microbiology Laboratory | Chan-Zuckerberg Biohub | CZB Cliahub Consortium et al |
| USA/CA-CDPH-UC10/2020 | EPI_ISL_413930 | 3/5/2020 | California Department of Public Health | Chiu Laboratory UCSF-Abbott Viral Diagnostics and Discovery Center University of California, San Francisco | Xianding Deng et al |
| USA/CA-UCSF-UC42/2020 | EPI_ISL_450234 | 3/14/2020 | UCSF Clinical Microbiology Laboratory | Chiu Laboratory, University of California, San Francisco | Xianding Deng et al |
| USA/CA-CZB-1111/2020 | EPI_ISL_444069 | 4/30/2020 | UCSF Clinical Microbiology Laboratory | Chan-Zuckerberg Biohub | CZB Cliahub Consortium et al |
| USA/CA-CZB-1126/2020 | EPI_ISL_445174 | 3/31/2020 | UCSF Clinical Microbiology Laboratory | Chan-Zuckerberg Biohub | CZB Cliahub Consortium et al |
| USA/CA-CZB-1122/2020 | EPI_ISL_445170 | 3/27/2020 | UCSF Clinical Microbiology Laboratory | Chan-Zuckerberg Biohub | CZB Cliahub Consortium et al |
| USA/CA-CZB0114/2020 | EPI_ISL_429058 | 3/28/2020 | UCSF Clinical Microbiology Laboratory | Chan-Zuckerberg Biohub | CZB Cliahub Consortium et al |
| USA/CA-CZB0141/2020 | EPI_ISL_429054 | 4/8/2020 | UCSF Clinical Microbiology Laboratory | Chan-Zuckerberg Biohub | CZB Cliahub Consortium et al |
| USA/CA-CZB017/2020 | EPI_ISL_429073 | 3/25/2020 | UCSF Clinical Microbiology Laboratory | Chan-Zuckerberg Biohub | CZB Cliahub Consortium et al |
| USA/CA-CZB-1124/2020 | EPI_ISL_445172 | 3/25/2020 | UCSF Clinical Microbiology Laboratory | Chan-Zuckerberg Biohub | CZB Cliahub Consortium et al |
| USA/CA-CZB-1125/2020 | EPI_ISL_445173 | 3/31/2020 | UCSF Clinical Microbiology Laboratory | Chan-Zuckerberg Biohub | CZB Cliahub Consortium et al |
| USA/CA-CZB0109/2020 | EPI_ISL_429066 | 3/28/2020 | UCSF Clinical Microbiology Laboratory | Chan-Zuckerberg Biohub | CZB Cliahub Consortium et al |
| USA/CA-CZB040/2020 | EPI_ISL_429064 | 3/25/2020 | UCSF Clinical Microbiology Laboratory | Chan-Zuckerberg Biohub | CZB Cliahub Consortium et al |
| USA/CA-CZB0108/2020 | EPI_ISL_429010 | 3/28/2020 | UCSF Clinical Microbiology Laboratory | Chan-Zuckerberg Biohub | CZB Cliahub Consortium et al |
| USA/CA-CZB0110/2020 | EPI_ISL_429059 | 3/28/2020 | UCSF Clinical Microbiology Laboratory | Chan-Zuckerberg Biohub | CZB Cliahub Consortium et al |
| USA/CA-CZB0133/2020 | EPI_ISL_428995 | 4/8/2020 | UCSF Clinical Microbiology Laboratory | Chan-Zuckerberg Biohub | CZB Cliahub Consortium et al |
| USA/CA-CZB-1093/2020 | EPI_ISL_444051 | 4/27/2020 | UCSF Clinical Microbiology Laboratory | Chan-Zuckerberg Biohub | CZB Cliahub Consortium et al |
| USA/CA-CZB-1097/2020 | EPI_ISL_444055 | 4/26/2020 | UCSF Clinical Microbiology Laboratory | Chan-Zuckerberg Biohub | CZB Cliahub Consortium et al |
| USA/CA-CZB0103/2020 | EPI_ISL_429000 | 3/28/2020 | UCSF Clinical Microbiology Laboratory | Chan-Zuckerberg Biohub | CZB Cliahub Consortium et al |
| USA/CA-CZB079/2020 | EPI_ISL_429005 | 3/28/2020 | UCSF Clinical Microbiology Laboratory | Chan-Zuckerberg Biohub | CZB Cliahub Consortium et al |
| USA/CA-CZB078/2020 | EPI_ISL_429019 | 3/28/2020 | UCSF Clinical Microbiology Laboratory | Chan-Zuckerberg Biohub | CZB Cliahub Consortium et al |
| USA/CA-CZB0701/2020 | EPI_ISL_429040 | 3/23/2020 | UCSF Clinical Microbiology Laboratory | Chan-Zuckerberg Biohub | CZB Cliahub Consortium et al |
| USA/CA-CZB045/2020 | EPI_ISL_429021 | 3/25/2020 | UCSF Clinical Microbiology Laboratory | Chan-Zuckerberg Biohub | CZB Cliahub Consortium et al |
| USA/CA-CZB091/2020 | EPI_ISL_429008 | 3/28/2020 | UCSF Clinical Microbiology Laboratory | Chan-Zuckerberg Biohub | CZB Cliahub Consortium et al |
| USA/CA-CZB0719/2020 | EPI_ISL_429072 | 3/31/2020 | UCSF Clinical Microbiology Laboratory | Chan-Zuckerberg Biohub | CZB Cliahub Consortium et al |
| USA/CA-CZB0725/2020 | EPI_ISL_429060 | 3/31/2020 | UCSF Clinical Microbiology Laboratory | Chan-Zuckerberg Biohub | CZB Cliahub Consortium et al |
| USA/CA-CZB04/2020 | EPI_ISL_429069 | 3/25/2020 | UCSF Clinical Microbiology Laboratory | Chan-Zuckerberg Biohub | CZB Cliahub Consortium et al |
| USA/CA-CZB0700/2020 | EPI_ISL_429070 | 3/23/2020 | UCSF Clinical Microbiology Laboratory | Chan-Zuckerberg Biohub | CZB Cliahub Consortium et al |
| USA/CA-CZB082/2020 | EPI_ISL_429032 | 3/28/2020 | UCSF Clinical Microbiology Laboratory | Chan-Zuckerberg Biohub | CZB Cliahub Consortium et al |
| USA/CA-CZB0746/2020 | EPI_ISL_429028 | 4/3/2020 | UCSF Clinical Microbiology Laboratory | Chan-Zuckerberg Biohub | CZB Cliahub Consortium et al |
| USA/CA-CZB0726/2020 | EPI_ISL_429055 | 3/31/2020 | UCSF Clinical Microbiology Laboratory | Chan-Zuckerberg Biohub | CZB Cliahub Consortium et al |
| USA/CA-CZB-1168/2020 | EPI_ISL_454622 | 4/27/2020 | UCSF Clinical Microbiology Laboratory | Chan-Zuckerberg Biohub | CZB Cliahub Consortium et al |
| USA/CA-CZB-1096/2020 | EPI_ISL_444054 | 4/29/2020 | UCSF Clinical Microbiology Laboratory | Chan-Zuckerberg Biohub | CZB Cliahub Consortium et al |
| USA/CA-CZB0158/2020 | EPI_ISL_429044 | 4/8/2020 | UCSF Clinical Microbiology Laboratory | Chan-Zuckerberg Biohub | CZB Cliahub Consortium et al |
| USA/CA-CZB027/2020 | EPI_ISL_429068 | 3/25/2020 | UCSF Clinical Microbiology Laboratory | Chan-Zuckerberg Biohub | CZB Cliahub Consortium et al |
| USA/CA-CZB-1104/2020 | EPI_ISL_444062 | 4/28/2020 | UCSF Clinical Microbiology Laboratory | Chan-Zuckerberg Biohub | CZB Cliahub Consortium et al |
| USA/CA-CDPH-UC28/2020 | EPI_ISL_417332 | 3/14/2020 | Chiu Laboratory, University of California, San Francisco | Chiu Laboratory, University of California, San Francisco | Xianding Deng et al |
| USA/CA-CZB096/2020 | EPI_ISL_429041 | 3/28/2020 | UCSF Clinical Microbiology Laboratory | Chan-Zuckerberg Biohub | CZB Cliahub Consortium et al |
| USA/CA-CZB-1107/2020 | EPI_ISL_444065 | 4/30/2020 | UCSF Clinical Microbiology Laboratory | Chan-Zuckerberg Biohub | CZB Cliahub Consortium et al |
| USA/CA-CZB-1157/2020 | EPI_ISL_454618 | 4/26/2020 | UCSF Clinical Microbiology Laboratory | Chan-Zuckerberg Biohub | CZB Cliahub Consortium et al |
| USA/CA-CZB-1153/2020 | EPI_ISL_454616 | 4/26/2020 | UCSF Clinical Microbiology Laboratory | Chan-Zuckerberg Biohub | CZB Cliahub Consortium et al |
| USA/CA-CZB090/2020 | EPI_ISL_429049 | 3/28/2020 | UCSF Clinical Microbiology Laboratory | Chan-Zuckerberg Biohub | CZB Cliahub Consortium et al |
| USA/CA-CZB0101/2020 | EPI_ISL_429056 | 3/28/2020 | UCSF Clinical Microbiology Laboratory | Chan-Zuckerberg Biohub | CZB Cliahub Consortium et al |
| USA/CA-CZB0112/2020 | EPI_ISL_429065 | 3/28/2020 | UCSF Clinical Microbiology Laboratory | Chan-Zuckerberg Biohub | CZB Cliahub Consortium et al |
| USA/CA-CZB0125/2020 | EPI_ISL_429046 | 4/8/2020 | UCSF Clinical Microbiology Laboratory | Chan-Zuckerberg Biohub | CZB Cliahub Consortium et al |
| USA/CA-CZB-1164/2020 | EPI_ISL_454621 | 4/27/2020 | UCSF Clinical Microbiology Laboratory | Chan-Zuckerberg Biohub | CZB Cliahub Consortium et al |
| USA/CA-CZB-1114/2020 | EPI_ISL_444072 | 4/30/2020 | UCSF Clinical Microbiology Laboratory | Chan-Zuckerberg Biohub | CZB Cliahub Consortium et al |
| USA/CA-CZB0737/2020 | EPI_ISL_429025 | 3/31/2020 | UCSF Clinical Microbiology Laboratory | Chan-Zuckerberg Biohub | CZB Cliahub Consortium et al |
| USA/CA-CZB036/2020 | EPI_ISL_429039 | 3/25/2020 | UCSF Clinical Microbiology Laboratory | Chan-Zuckerberg Biohub | CZB Cliahub Consortium et al |
| USA/CA-CZB015b/2020 | EPI_ISL_428997 | 3/25/2020 | UCSF Clinical Microbiology Laboratory | Chan-Zuckerberg Biohub | CZB Cliahub Consortium et al |
| USA/CA-CZB039/2020 | EPI_ISL_429045 | 3/25/2020 | UCSF Clinical Microbiology Laboratory | Chan-Zuckerberg Biohub | CZB Cliahub Consortium et al |
| USA/CA-CZB016/2020 | EPI_ISL_429012 | 3/25/2020 | UCSF Clinical Microbiology Laboratory | Chan-Zuckerberg Biohub | CZB Cliahub Consortium et al |
| USA/CA-UCSF-UC43/2020 | EPI_ISL_450235 | 3/14/2020 | UCSF Clinical Microbiology Laboratory | Chiu Laboratory, University of California, San Francisco | Xianding Deng et al |
| USA/CA-CZB0716/2020 | EPI_ISL_430792 | 3/23/2020 | UCSF Clinical Microbiology Laboratory | Chan-Zuckerberg Biohub | CZB Cliahub Consortium et al |
| USA/CA-CZB088/2020 | EPI_ISL_430791 | 3/28/2020 | UCSF Clinical Microbiology Laboratory | Chan-Zuckerberg Biohub | CZB Cliahub Consortium et al |
| USA/CA-CZB095/2020 | EPI_ISL_429006 | 3/28/2020 | UCSF Clinical Microbiology Laboratory | Chan-Zuckerberg Biohub | CZB Cliahub Consortium et al |
| USA/CA-CZB-1110/2020 | EPI_ISL_444068 | 4/30/2020 | UCSF Clinical Microbiology Laboratory | Chan-Zuckerberg Biohub | CZB Cliahub Consortium et al |
| USA/CA-CZB-1098/2020 | EPI_ISL_444056 | 4/26/2020 | UCSF Clinical Microbiology Laboratory | Chan-Zuckerberg Biohub | CZB Cliahub Consortium et al |
| USA/CA-CZB-1170/2020 | EPI_ISL_454623 | 4/29/2020 | UCSF Clinical Microbiology Laboratory | Chan-Zuckerberg Biohub | CZB Cliahub Consortium et al |
| USA/CA-CZB-1175/2020 | EPI_ISL_454626 | 4/30/2020 | UCSF Clinical Microbiology Laboratory | Chan-Zuckerberg Biohub | CZB Cliahub Consortium et al |
| USA/CA-UCSF-UC33/2020 | EPI_ISL_429878 | 3/7/2020 | Chiu Laboratory, University of California, San Francisco | Chiu Laboratory, University of California, San Francisco | Xianding Deng et al |
| USA/CA-UCSF-UC32/2020 | EPI_ISL_429877 | 3/5/2020 | Chiu Laboratory, University of California, San Francisco | Chiu Laboratory, University of California, San Francisco | Xianding Deng et al |
| USA/CA-CZB046/2020 | EPI_ISL_429001 | 3/25/2020 | UCSF Clinical Microbiology Laboratory | Chan-Zuckerberg Biohub | CZB Cliahub Consortium et al |
| USA/CA-CZB0706/2020 | EPI_ISL_429034 | 3/23/2020 | UCSF Clinical Microbiology Laboratory | Chan-Zuckerberg Biohub | CZB Cliahub Consortium et al |
| USA/CA-CZB0458/2020 | EPI_ISL_429007 | 3/18/2020 | UCSF Clinical Microbiology Laboratory | Chan-Zuckerberg Biohub | CZB Cliahub Consortium et al |
| USA/CA-CZB0107/2020 | EPI_ISL_429036 | 3/28/2020 | UCSF Clinical Microbiology Laboratory | Chan-Zuckerberg Biohub | CZB Cliahub Consortium et al |
| USA/CA-CZB0145/2020 | EPI_ISL_429062 | 4/8/2020 | UCSF Clinical Microbiology Laboratory | Chan-Zuckerberg Biohub | CZB Cliahub Consortium et al |
| USA/CA-CZB05/2020 | EPI_ISL_417931 | 3/18/2020 | UCSF Clinical Microbiology Laboratory | Chan-Zuckerberg Biohub | Shaun Arevalo et al |
| USA/CA-UCSF-UC41/2020 | EPI_ISL_450233 | 3/14/2020 | UCSF Clinical Microbiology Laboratory | Chiu Laboratory, University of California, San Francisco | Xianding Deng et al |
| USA/CA-CZB089/2020 | EPI_ISL_429050 | 3/28/2020 | UCSF Clinical Microbiology Laboratory | Chan-Zuckerberg Biohub | CZB Cliahub Consortium et al |
| USA/CA-CZB085/2020 | EPI_ISL_429004 | 3/28/2020 | UCSF Clinical Microbiology Laboratory | Chan-Zuckerberg Biohub | CZB Cliahub Consortium et al |
| USA/CA-UCSF-UC47/2020 | EPI_ISL_450239 | 3/20/2020 | UCSF Clinical Microbiology Laboratory | Chiu Laboratory, University of California, San Francisco | Xianding Deng et al |
| USA/CA-CZB067/2020 | EPI_ISL_429029 | 3/28/2020 | UCSF Clinical Microbiology Laboratory | Chan-Zuckerberg Biohub | CZB Cliahub Consortium et al |
| USA/CA-CZB-1127/2020 | EPI_ISL_445175 | 4/5/2020 | UCSF Clinical Microbiology Laboratory | Chan-Zuckerberg Biohub | CZB Cliahub Consortium et al |
| USA/CA-CZB-1128/2020 | EPI_ISL_445176 | 4/5/2020 | UCSF Clinical Microbiology Laboratory | Chan-Zuckerberg Biohub | CZB Cliahub Consortium et al |
| USA/CA-CZB0707/2020 | EPI_ISL_429038 | 3/23/2020 | UCSF Clinical Microbiology Laboratory | Chan-Zuckerberg Biohub | CZB Cliahub Consortium et al |
| USA/CA-UCSF-UC45/2020 | EPI_ISL_450237 | 3/19/2020 | UCSF Clinical Microbiology Laboratory | Chiu Laboratory, University of California, San Francisco | Xianding Deng et al |
| USA/CA-CZB0703/2020 | EPI_ISL_428994 | 3/23/2020 | UCSF Clinical Microbiology Laboratory | Chan-Zuckerberg Biohub | CZB Cliahub Consortium et al |
| USA/CA-CZB-1186/2020 | EPI_ISL_454631 | 4/29/2020 | UCSF Clinical Microbiology Laboratory | Chan-Zuckerberg Biohub | CZB Cliahub Consortium et al |
| USA/CA-CZB0167/2020 | EPI_ISL_429053 | 4/8/2020 | UCSF Clinical Microbiology Laboratory | Chan-Zuckerberg Biohub | CZB Cliahub Consortium et al |
| USA/CA-CZB0713/2020 | EPI_ISL_429022 | 3/23/2020 | UCSF Clinical Microbiology Laboratory | Chan-Zuckerberg Biohub | CZB Cliahub Consortium et al |
| USA/CA-CZB07/2020 | EPI_ISL_417933 | 3/18/2020 | UCSF Clinical Microbiology Laboratory | Chan-Zuckerberg Biohub | Shaun Arevalo et al |
| USA/CA-UCSF-UC40/2020 | EPI_ISL_450232 | 3/15/2020 | UCSF Clinical Microbiology Laboratory | Chiu Laboratory, University of California, San Francisco | Xianding Deng et al |
| USA/CA-CZB0714/2020 | EPI_ISL_429052 | 3/23/2020 | UCSF Clinical Microbiology Laboratory | Chan-Zuckerberg Biohub | CZB Cliahub Consortium et al |
| USA/CA-CZB-1123/2020 | EPI_ISL_445171 | 3/27/2020 | UCSF Clinical Microbiology Laboratory | Chan-Zuckerberg Biohub | CZB Cliahub Consortium et al |
| USA/CA-UCSF-UC46/2020 | EPI_ISL_450238 | 3/17/2020 | UCSF Clinical Microbiology Laboratory | Chiu Laboratory, University of California, San Francisco | Xianding Deng et al |
| USA/CA-UCSF-UC36/2020 | EPI_ISL_429881 | 3/20/2020 | California Department of Public Health | Chiu Laboratory, University of California, San Francisco | Xianding Deng et al |
| USA/CA-CZB-1193/2020 | EPI_ISL_454633 | 4/29/2020 | UCSF Clinical Microbiology Laboratory | Chan-Zuckerberg Biohub | CZB Cliahub Consortium et al |
| USA/CA-CZB-1174/2020 | EPI_ISL_454625 | 4/30/2020 | UCSF Clinical Microbiology Laboratory | Chan-Zuckerberg Biohub | CZB Cliahub Consortium et al |
| USA/CA-CZB-1106/2020 | EPI_ISL_444064 | 4/30/2020 | UCSF Clinical Microbiology Laboratory | Chan-Zuckerberg Biohub | CZB Cliahub Consortium et al |
| USA/CA-CZB-1118/2020 | EPI_ISL_444076 | 4/30/2020 | UCSF Clinical Microbiology Laboratory | Chan-Zuckerberg Biohub | CZB Cliahub Consortium et al |
| USA/CA-CZB-1117/2020 | EPI_ISL_444075 | 4/30/2020 | UCSF Clinical Microbiology Laboratory | Chan-Zuckerberg Biohub | CZB Cliahub Consortium et al |
| USA/CA-CZB-1120/2020 | EPI_ISL_444078 | 4/29/2020 | UCSF Clinical Microbiology Laboratory | Chan-Zuckerberg Biohub | CZB Cliahub Consortium et al |
| USA/CA-CZB-1108/2020 | EPI_ISL_444066 | 4/30/2020 | UCSF Clinical Microbiology Laboratory | Chan-Zuckerberg Biohub | CZB Cliahub Consortium et al |
| USA/CA-CZB-1105/2020 | EPI_ISL_444063 | 4/30/2020 | UCSF Clinical Microbiology Laboratory | Chan-Zuckerberg Biohub | CZB Cliahub Consortium et al |
| USA/CA-CZB-1146/2020 | EPI_ISL_454614 | 4/29/2020 | UCSF Clinical Microbiology Laboratory | Chan-Zuckerberg Biohub | CZB Cliahub Consortium et al |
| USA/CA-CZB-1119/2020 | EPI_ISL_444077 | 4/30/2020 | UCSF Clinical Microbiology Laboratory | Chan-Zuckerberg Biohub | CZB Cliahub Consortium et al |
| USA/CA-CZB-1109/2020 | EPI_ISL_444067 | 4/30/2020 | UCSF Clinical Microbiology Laboratory | Chan-Zuckerberg Biohub | CZB Cliahub Consortium et al |
| USA/CA-CZB-1103/2020 | EPI_ISL_444061 | 4/27/2020 | UCSF Clinical Microbiology Laboratory | Chan-Zuckerberg Biohub | CZB Cliahub Consortium et al |
| USA/CA-CZB-1095/2020 | EPI_ISL_444053 | 4/27/2020 | UCSF Clinical Microbiology Laboratory | Chan-Zuckerberg Biohub | CZB Cliahub Consortium et al |
| USA/CA-CZB-1094/2020 | EPI_ISL_444052 | 4/27/2020 | UCSF Clinical Microbiology Laboratory | Chan-Zuckerberg Biohub | CZB Cliahub Consortium et al |
| USA/CA-CZB-1101/2020 | EPI_ISL_444059 | 4/27/2020 | UCSF Clinical Microbiology Laboratory | Chan-Zuckerberg Biohub | CZB Cliahub Consortium et al |
| USA/CA-CZB-1102/2020 | EPI_ISL_444060 | 4/27/2020 | UCSF Clinical Microbiology Laboratory | Chan-Zuckerberg Biohub | CZB Cliahub Consortium et al |
| USA/CA-CZB030/2020 | EPI_ISL_429017 | 3/25/2020 | UCSF Clinical Microbiology Laboratory | Chan-Zuckerberg Biohub | CZB Cliahub Consortium et al |
| USA/CA-CZB080/2020 | EPI_ISL_429013 | 3/28/2020 | UCSF Clinical Microbiology Laboratory | Chan-Zuckerberg Biohub | CZB Cliahub Consortium et al |
| USA/CA-CZB0100/2020 | EPI_ISL_429015 | 3/28/2020 | UCSF Clinical Microbiology Laboratory | Chan-Zuckerberg Biohub | CZB Cliahub Consortium et al |
| USA/CA-CZB0709/2020 | EPI_ISL_429016 | 3/23/2020 | UCSF Clinical Microbiology Laboratory | Chan-Zuckerberg Biohub | CZB Cliahub Consortium et al |
| USA/CA-CZB0731/2020 | EPI_ISL_429026 | 3/31/2020 | UCSF Clinical Microbiology Laboratory | Chan-Zuckerberg Biohub | CZB Cliahub Consortium et al |
| USA/CA-CZB-1132/2020 | EPI_ISL_445180 | 3/30/2020 | UCSF Clinical Microbiology Laboratory | Chan-Zuckerberg Biohub | CZB Cliahub Consortium et al |
| USA/CA-CZB0147/2020 | EPI_ISL_429011 | 4/8/2020 | UCSF Clinical Microbiology Laboratory | Chan-Zuckerberg Biohub | CZB Cliahub Consortium et al |
| USA/CA-CZB0740/2020 | EPI_ISL_429023 | 3/31/2020 | UCSF Clinical Microbiology Laboratory | Chan-Zuckerberg Biohub | CZB Cliahub Consortium et al |
| USA/CA-CZB0750/2020 | EPI_ISL_429018 | 4/3/2020 | UCSF Clinical Microbiology Laboratory | Chan-Zuckerberg Biohub | CZB Cliahub Consortium et al |
| USA/CA-CDPH-UC27/2020 | EPI_ISL_417331 | 3/13/2020 | Chiu Laboratory, University of California, San Francisco | Chiu Laboratory, University of California, San Francisco | Xianding Deng et al |
| USA/CA-UCSF-UC48/2020 | EPI_ISL_450240 | 3/20/2020 | UCSF Clinical Microbiology Laboratory | Chiu Laboratory, University of California, San Francisco | Xianding Deng et al |
| USA/CA-CZB013/2020 | EPI_ISL_417937 | 3/18/2020 | UCSF Clinical Microbiology Laboratory | Chan-Zuckerberg Biohub | Shaun Arevalo et al |
| USA/CA-CZB0741/2020 | EPI_ISL_429051 | 4/3/2020 | UCSF Clinical Microbiology Laboratory | Chan-Zuckerberg Biohub | CZB Cliahub Consortium et al |
| USA/CA-CZB084/2020 | EPI_ISL_428998 | 3/28/2020 | UCSF Clinical Microbiology Laboratory | Chan-Zuckerberg Biohub | CZB Cliahub Consortium et al |
| USA/CA-CZB-1134/2020 | EPI_ISL_445182 | 4/12/2020 | UCSF Clinical Microbiology Laboratory | Chan-Zuckerberg Biohub | CZB Cliahub Consortium et al |
| USA/CA-CZB-1195/2020 | EPI_ISL_454634 | 4/29/2020 | UCSF Clinical Microbiology Laboratory | Chan-Zuckerberg Biohub | CZB Cliahub Consortium et al |
| USA/CA-CZB-1113/2020 | EPI_ISL_444071 | 4/30/2020 | UCSF Clinical Microbiology Laboratory | Chan-Zuckerberg Biohub | CZB Cliahub Consortium et al |
| USA/CA-CZB09/2020 | EPI_ISL_429037 | 3/25/2020 | UCSF Clinical Microbiology Laboratory | Chan-Zuckerberg Biohub | CZB Cliahub Consortium et al |
| USA/CA-CZB0710/2020 | EPI_ISL_429042 | 3/23/2020 | UCSF Clinical Microbiology Laboratory | Chan-Zuckerberg Biohub | CZB Cliahub Consortium et al |
| USA/CA-CZB-1133/2020 | EPI_ISL_445181 | 4/8/2020 | UCSF Clinical Microbiology Laboratory | Chan-Zuckerberg Biohub | CZB Cliahub Consortium et al |
| USA/CA-CZB0138/2020 | EPI_ISL_428992 | 4/8/2020 | UCSF Clinical Microbiology Laboratory | Chan-Zuckerberg Biohub | CZB Cliahub Consortium et al |
| USA/CA-CZB06/2020 | EPI_ISL_417932 | 3/18/2020 | UCSF Clinical Microbiology Laboratory | Chan-Zuckerberg Biohub | Shaun Arevalo et al |
| USA/CA-CZB0137/2020 | EPI_ISL_429027 | 4/8/2020 | UCSF Clinical Microbiology Laboratory | Chan-Zuckerberg Biohub | CZB Cliahub Consortium et al |
| USA/CA-CZB0136/2020 | EPI_ISL_429003 | 4/8/2020 | UCSF Clinical Microbiology Laboratory | Chan-Zuckerberg Biohub | CZB Cliahub Consortium et al |
| USA/CA-CZB0122/2020 | EPI_ISL_429009 | 4/8/2020 | UCSF Clinical Microbiology Laboratory | Chan-Zuckerberg Biohub | CZB Cliahub Consortium et al |
| USA/CA-CZB-1131/2020 | EPI_ISL_445179 | 3/31/2020 | UCSF Clinical Microbiology Laboratory | Chan-Zuckerberg Biohub | CZB Cliahub Consortium et al |
| USA/CA-CZB-1121/2020 | EPI_ISL_445169 | 3/28/2020 | UCSF Clinical Microbiology Laboratory | Chan-Zuckerberg Biohub | CZB Cliahub Consortium et al |
| USA/CA-CZB071/2020 | EPI_ISL_428991 | 3/28/2020 | UCSF Clinical Microbiology Laboratory | Chan-Zuckerberg Biohub | CZB Cliahub Consortium et al |
| USA/CA-CZB015a/2020 | EPI_ISL_417939 | 3/18/2020 | UCSF Clinical Microbiology Laboratory | Chan-Zuckerberg Biohub | Shaun Arevalo et al |
| USA/CA-CZB014/2020 | EPI_ISL_417938 | 3/18/2020 | UCSF Clinical Microbiology Laboratory | Chan-Zuckerberg Biohub | Shaun Arevalo et al |
| USA/CA-CZB0156/2020 | EPI_ISL_429043 | 4/8/2020 | UCSF Clinical Microbiology Laboratory | Chan-Zuckerberg Biohub | CZB Cliahub Consortium et al |
| USA/CA-CZB0139/2020 | EPI_ISL_429031 | 4/8/2020 | UCSF Clinical Microbiology Laboratory | Chan-Zuckerberg Biohub | CZB Cliahub Consortium et al |
| USA/CA-CZB0142/2020 | EPI_ISL_429014 | 4/8/2020 | UCSF Clinical Microbiology Laboratory | Chan-Zuckerberg Biohub | CZB Cliahub Consortium et al |
| USA/CA-CZB-1189/2020 | EPI_ISL_454632 | 4/29/2020 | UCSF Clinical Microbiology Laboratory | Chan-Zuckerberg Biohub | CZB Cliahub Consortium et al |
| USA/CA-CZB-1116/2020 | EPI_ISL_444074 | 4/30/2020 | UCSF Clinical Microbiology Laboratory | Chan-Zuckerberg Biohub | CZB Cliahub Consortium et al |
| USA/CA-CZB-1115/2020 | EPI_ISL_444073 | 4/30/2020 | UCSF Clinical Microbiology Laboratory | Chan-Zuckerberg Biohub | CZB Cliahub Consortium et al |
| USA/CA-UCSF-UC44/2020 | EPI_ISL_450236 | 3/19/2020 | UCSF Clinical Microbiology Laboratory | Chiu Laboratory, University of California, San Francisco | Xianding Deng et al |
| USA/CA-CZB0705/2020 | EPI_ISL_429024 | 3/23/2020 | UCSF Clinical Microbiology Laboratory | Chan-Zuckerberg Biohub | CZB Cliahub Consortium et al |
| USA/CA-CDPH-UC26/2020 | EPI_ISL_417330 | 3/13/2020 | Chiu Laboratory, University of California, San Francisco | Chiu Laboratory, University of California, San Francisco | Xianding Deng et al |
| USA/CA-CZB0157/2020 | EPI_ISL_428993 | 4/8/2020 | UCSF Clinical Microbiology Laboratory | Chan-Zuckerberg Biohub | CZB Cliahub Consortium et al |
| USA/CA-CZB-1130/2020 | EPI_ISL_445178 | 4/5/2020 | UCSF Clinical Microbiology Laboratory | Chan-Zuckerberg Biohub | CZB Cliahub Consortium et al |
| USA/CA-CZB086/2020 | EPI_ISL_429033 | 3/28/2020 | UCSF Clinical Microbiology Laboratory | Chan-Zuckerberg Biohub | CZB Cliahub Consortium et al |
| USA/CA-CZB072/2020 | EPI_ISL_429030 | 3/28/2020 | UCSF Clinical Microbiology Laboratory | Chan-Zuckerberg Biohub | CZB Cliahub Consortium et al |
| USA/CA-CZB042/2020 | EPI_ISL_429035 | 3/25/2020 | UCSF Clinical Microbiology Laboratory | Chan-Zuckerberg Biohub | CZB Cliahub Consortium et al |
| USA/CA-CZB011a/2020 | EPI_ISL_417935 | 3/18/2020 | UCSF Clinical Microbiology Laboratory | Chan-Zuckerberg Biohub | Shaun Arevalo et al |
| USA/CA-CZB-1129/2020 | EPI_ISL_445177 | 3/31/2020 | UCSF Clinical Microbiology Laboratory | Chan-Zuckerberg Biohub | CZB Cliahub Consortium et al |
| USA/CA-CZB-1150/2020 | EPI_ISL_454615 | 4/26/2020 | UCSF Clinical Microbiology Laboratory | Chan-Zuckerberg Biohub | CZB Cliahub Consortium et al |
| USA/CA-CZB-1160/2020 | EPI_ISL_454619 | 4/26/2020 | UCSF Clinical Microbiology Laboratory | Chan-Zuckerberg Biohub | CZB Cliahub Consortium et al |
| USA/CA-CZB098/2020 | EPI_ISL_429002 | 3/28/2020 | UCSF Clinical Microbiology Laboratory | Chan-Zuckerberg Biohub | CZB Cliahub Consortium et al |
| USA/CA-CZB0104/2020 | EPI_ISL_428990 | 3/28/2020 | UCSF Clinical Microbiology Laboratory | Chan-Zuckerberg Biohub | CZB Cliahub Consortium et al |
| USA/CA-CZB-1099/2020 | EPI_ISL_444057 | 4/28/2020 | UCSF Clinical Microbiology Laboratory | Chan-Zuckerberg Biohub | CZB Cliahub Consortium et al |
| USA/CA-CZB-1112/2020 | EPI_ISL_444070 | 4/30/2020 | UCSF Clinical Microbiology Laboratory | Chan-Zuckerberg Biohub | CZB Cliahub Consortium et al |
| USA/CA-CZB-1163/2020 | EPI_ISL_454620 | 4/27/2020 | UCSF Clinical Microbiology Laboratory | Chan-Zuckerberg Biohub | CZB Cliahub Consortium et al |
| USA/CA-CZB0735/2020 | EPI_ISL_429067 | 3/31/2020 | UCSF Clinical Microbiology Laboratory | Chan-Zuckerberg Biohub | CZB Cliahub Consortium et al |
| USA/CA-CZB077/2020 | EPI_ISL_428996 | 3/28/2020 | UCSF Clinical Microbiology Laboratory | Chan-Zuckerberg Biohub | CZB Cliahub Consortium et al |
| USA/CA-CZB-1178/2020 | EPI_ISL_454628 | 4/30/2020 | UCSF Clinical Microbiology Laboratory | Chan-Zuckerberg Biohub | CZB Cliahub Consortium et al |
| USA/CA-CZB-1183/2020 | EPI_ISL_454630 | 4/30/2020 | UCSF Clinical Microbiology Laboratory | Chan-Zuckerberg Biohub | CZB Cliahub Consortium et al |
| USA/CA-CZB-1156/2020 | EPI_ISL_454617 | 4/26/2020 | UCSF Clinical Microbiology Laboratory | Chan-Zuckerberg Biohub | CZB Cliahub Consortium et al |
| USA/CA-CZB-1176/2020 | EPI_ISL_454627 | 4/30/2020 | UCSF Clinical Microbiology Laboratory | Chan-Zuckerberg Biohub | CZB Cliahub Consortium et al |
| USA/CA-CZB-1179/2020 | EPI_ISL_454629 | 4/30/2020 | UCSF Clinical Microbiology Laboratory | Chan-Zuckerberg Biohub | CZB Cliahub Consortium et al |
| USA/CA-CZB-1171/2020 | EPI_ISL_454624 | 4/30/2020 | UCSF Clinical Microbiology Laboratory | Chan-Zuckerberg Biohub | CZB Cliahub Consortium et al |
| USA/CA-CZB069/2020 | EPI_ISL_428999 | 3/28/2020 | UCSF Clinical Microbiology Laboratory | Chan-Zuckerberg Biohub | CZB Cliahub Consortium et al |

**Figure S1:**
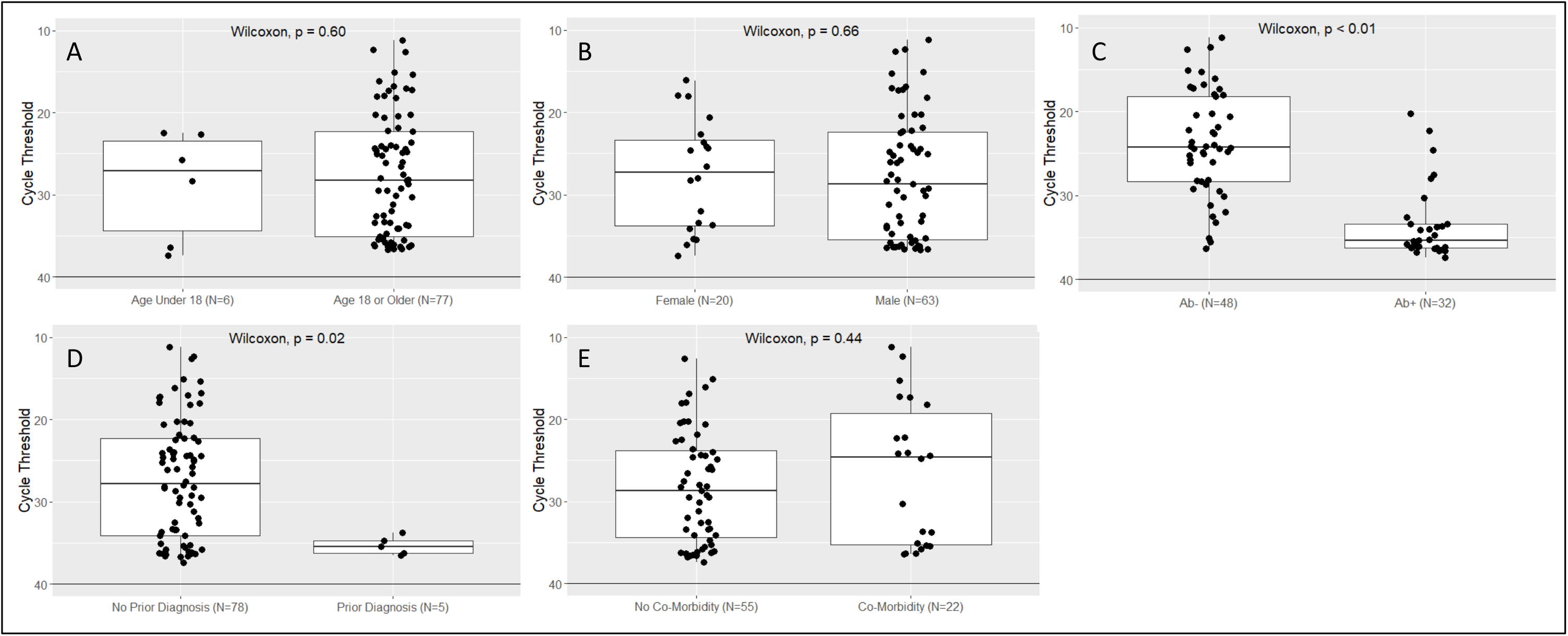
Levels of virus as estimated by RT-PCR cycle thresholds stratified by sub-group. RT-PCR cycle thresholds by age <18 vs. ≥18 years (Panel A), sex (Panel B), SARS-CoV-2 antibody status (Panel C), reported prior COVID-19 diagnosis (Panel D), and reported underlying medical conditions (Panel E) among PCR-positive participants (N=83).

1 1827 true negatives (18 true negatives) would represent a true recent prevalence of 1% in Bolinas. Even if sensitivity is 30% and specificity is 100%, the probability of zero test positives in Bolinas given 18 true positives is less than 0.2%. If sensitivity is higher or specificity is lower, the probability of zero test positives in Bolinas is even lower.

2 In the package insert it is noted: “Five specimens from 1 immunocompromised patient were excluded from the study. … When the results from these specimens were included, the PPA at ≥ 14 days post-symptom onset was 96.77% (95% CI: 90.86, 99.33).”

## Notes

### Competing Interest Statement

Mary Rodgers and John Hackett Jr. work for Abbott Laboratories. ARCHITECT SARS-CoV-2 test kits were provided by Abbott Laboratories. Charles Chiu is the director of the UCSF-Abbott Viral Diagnostics and Discovery Center (VDDC) and receives research support funding from Abbott Laboratories.

### Author Declarations

The University of California, San Francisco (UCSF) Committee on Human Research determined that the study met criteria for public health surveillance.

